# Slowing the spread of treatment failure to artemisinin-based combination therapies in Uganda

**DOI:** 10.1101/2025.05.15.25327701

**Authors:** Tran Dang Nguyen, Robert J Zupko, Melissa D Conrad, Gerald B Rukundo, Carter C Farinha, Victor D Asua, Kien Trung Tran, Deborah M Grace, Philip J Rosenthal, Bosco B Agaba, Moses R Kamya, Jimmy Opigo, Maciej F Boni

## Abstract

**Background:** The multiple emergences and continuing spread of partially artemisinin-resistant *Plasmodium falciparum* in Africa, where about 95% of malaria occurs, is a health challenge that requires urgent attention. The World Health Organization has developed a resistance response strategy that centers on enhancing surveillance, reducing drug pressure, and evaluating novel tools to slow resistance evolution which includes the deployment of multiple first-line therapies (MFT). Developing a specific resistance response is critical for Uganda, where four *pfkelch13* mutations are at local allele frequencies >0.20.

**Methods:** Using a previously validated Uganda-calibrated individual-based mathematical model of *P. falciparum* transmission and evolution, we evaluated 53 public-sector deployment strategies for artemisinin-based combination therapies (ACTs) aimed at reducing treatment failure and slowing the spread of *pfkelch13* alleles from 2025 to 2031. We assume that artemether-lumefantrine (AL) will continue to be used in the private sector.

**Results:** A change of first-line therapy from AL to artesunate-amodiaquine (ASAQ) is projected to reduce treatment failures by 34.7% to 38.3% (90% range of simulation outcomes) over six years, while a change to dihydroartemisinin-piperaquine (DHA-PPQ) is projected to reduce treatment failures over the same period by 10.0% to 12.9%. This pessimistic projection for DHA-PPQ deployment rests on a model assumption – supported by clinical data from SE Asia – that piperaquine resistance evolution will lead to high rates of treatment failure. Optimal MFT deployments and cycling approaches are projected to reduce treatment failure counts by ∼36% when compared to status quo AL use, an outcome similar to country-wide ASAQ deployment. MFT and cycling approaches are predicted to work best when ASAQ is recommended for a majority of malaria cases and DHA-PPQ for a smaller proportion of cases. Deployment of the triple ACT artemether-lumefantrine-amodiaquine has the potential to reduce treatment failures by ∼42% if enacted immediately.

**Conclusions:** Increased adoption of and coverage with ASAQ is projected to play a large role in reducing malaria treatment failure counts in Uganda over the next six years. With continued AL use in the private sector, ASAQ and DHA-PPQ deployment in the public sector creates a public-private MFT mix of antimalarial use. DHA-PPQ deployment should be accompanied by real-time molecular surveillance for piperaquine-resistant genotypes.

## 1 Introduction

The 2024 World Malaria Report indicates a slowly increasing global annual case burden, with 263 million malaria episodes and 597,000 malaria deaths estimated for 2023 [1]. Across multiple malaria management challenges, one clear and present concern is the spread of artemisinin partial resistance in *Plasmodium falciparum* in Africa. These resistant *P. falciparum* genotypes carry mutations in the *pfkelch13* gene. A total of six validated mutations – i.e. mutations that are associated with longer parasite clearance times and reduced artemisinin susceptibility in vitro [2] – have been identified across eight countries in east and central Africa [3–5]. This presents a clear urgency to respond to and prepare for the further spread of artemisinin resistance across the continent [6–8]. Global guidelines for the response are being formulated [9,10] and some national-level response planning is underway [11]. Custom response and preparation strategies will need to be developed for each country, as partner-drug landscapes, drug markets, health systems, and malaria transmission levels differ.

Until new non-artemisinin antimalarial therapies become available, one approach to slowing *pfkelch13* evolution – thus, slowing the spread of resistance and reducing treatment failures – will involve diversification of the partner drugs contained in ACTs [9,12]. The three primary ACTs deployed in this diversification will probably be artemether-lumefantrine (AL), artesunate-amodiaquine (ASAQ), and dihydroartemisinin-piperaquine (DHA-PPQ). AL is the most widely used ACT, but it will likely have its usage rate reduced in many settings due to nearly twenty years of consistent use that has possibly led to reduced levels of *P. falciparum* sensitivity to the partner-drug lumefantrine. A fourth ACT that is being considered for increased procurement and accessibility is artesunate-pyronaridine (ASPY) [13,14] which has been shown in therapeutic efficacy studies (TES) to have high efficacy in Africa [15–17]. At the moment, it is not known what the manufacturing and procurement capacity of ASPY will be in the coming years in East Africa.

Among epidemiological settings in Africa where the spread of kelch variants (falciparum parasites that carry validated partially artemisinin-resistant *pfkelch13* mutations) has been documented, Uganda’s situation is arguably the most dire with four kelch variants circulating at allele frequencies >0.20 in some districts and several others detected at lower frequencies [18]. Uganda also falls into a group of high-transmission countries where high case numbers and a low detection rate of new infections make many aspects of malaria control challenging. Nevertheless, the Ugandan National Malaria Elimination Division (NMED) began planning artemisinin-resistance response strategies in 2023, with a focus on changing drug policy. Here, we evaluate the first candidate set of approaches that should be discussed in any African context facing spreading artemisinin resistance, specifically changes to first-line therapy, introduction of ACT cycling regimens, deployment of multiple first-line therapies (MFT), and/or deployment of triple artemisinin-combination therapies.

## 2 Methods

### 2.1 Geography, Population, Travel Time

As described previously [11,19], a spatial individual-based *Plasmodium falciparum* transmission and evolution model was calibrated to Uganda’s geography, population, malaria prevalence, and malaria treatment patterns. Shapefiles (national and subnational boundaries) for Uganda were downloaded from the Humanitarian Data Exchange [20] along with the 2018 Malaria Indicator Survey (MIS) boundaries [21], and converted to the appropriate file type and re-projected as necessary to WGS 84 / UTM zone 36N (WKID 32636). Since the simulation runs in a spatial configuration based on the raster data, misalignments between the subnational boundaries and the MIS boundaries were not addressed, and the subnational boundaries were used as the canonical source when preparing the raster files for the simulation. Individual mortality and population demographic data were based upon the United Nations World Population Prospects (UN WPP) reports from 2019.

Following preparation of the shapefiles, raster data were next augmented by acquiring the 2020 population density data from WorldPop [22] which were then re-projected and aggregated to 25 km^2^ pixels from 30 arc-second pixels, with rounding up to the nearest whole number. This resulted in a total of 8,566 grid points, which were assigned to one of fifteen MIS districts based upon the 2018 MIS boundaries [21]. The aggregated data were then clipped to match the regional boundaries defined above. A similar process was used for the travel friction surface, which was acquired from the Malaria Atlas Project’s map of travel time in minutes to the nearest city, for the year 2015 [23]. The travel friction surface is used to augment a gravity model of travel in sub-Saharan Africa [24], by biasing individuals’ travel to the nearest large city as opposed to the largest city in the country [25] (Supp Tables S4, S5, S6).

### 2.2 Calibration of Prevalence and Treatment Seeking Patterns

Treatment seeking data were obtained from the 2018 MIS and assigned to the fifteen MIS regions in Table 1. These data were collected from December 11 2018 to January 31 2019 for children under five who had a fever in the preceding two weeks; children were counted as “treated” if advice or treatment was sought. The Kampala region did not receive enough survey responses for a reliable estimate.

**Table 1.** Treatment Coverage by MIS region. The Uganda 2018-2019 MIS contained data for fourteen of the fifteen MIS regions. The Kampala region recorded fewer than 25 responses, and the coverage estimate used in the simulation was 93%, which is the average of treatment coverages from bordering regions.

| MIS Region | Treatment Coverage for children under 5 |
| --- | --- |
| West Nile | 86% |
| Acholi | 87% |
| Karamoja | 85% |
| Bunyoro | 85% |
| Lango | 95% |
| Teso | 81% |
| Bugisu | 85% |
| Tooro | 79% |
| South Buganda | 97% |
| Busoga | 88% |
| Bukedi | 81% |
| Kigezi | 88% |
| Ankole | 91% |
| North Buganda | 89% |
| Kampala | insufficient response rate in survey |

In order to account for seasonal fluctuations in *Anopheles* vector abundance, rainfall data were used as a proxy to adjust the daily transmission rate. Eleven years (2009-01-01 to 2019-12-31) of daily rainfall data within Uganda’s national borders from ERA5 [26] were aggregated using Google Earth Engine. The daily means were then calculated, smoothed, and shifted forward by eleven days to account for the Anopheles lifecycle. This annual pattern was used to create an approximation of temporal variation in biting patterns as seen in Supp Figure S1.

Following the collection of data on treatment seeking, drug choice, and rainfall patterns (along with the preparation of the population, district, and travel rasters for the model) we calibrated each pixel’s generic transmission parameter or biting parameter (β), which controls the transmission intensity of *P. falciparum* in the simulation. The calibration aims to match, for each pixel, the model’s 12-month average of *P. falciparum* prevalence in 2-10 year-olds (PfPR_2-10_) to Malaria Atlas Project estimates of the same quantity for 2017 [27], as described previously [11,19,28]. Once complete, these MAP-informed pixel prevalences were updated to better match 2018-2019 PfPR_0-59_ estimates (prevalence in children aged zero to 59 months) present in the 2018-2019 MIS (see Supp Table S2). However, this approach introduces some artifacts into the projected prevalence due to the Malaria Indicator Survey 2018-2019 survey district prevalence data being weighted higher, resulting in some spatial prevalence distributions that follow political boundaries.

### 2.3 Incidence Calibration

In addition to verifying that the PfPR_0-59_ was within ±1% (in absolute terms, for 14 of 15 MIS regions) of the estimates from the 2018 MIS, the clinical case counts per 1000 individuals were also checked against the target calibration year of 2020. In the simulation, all clinical cases (i.e. symptomatic episodes of malaria) were logged along with their treatment or lack thereof; not all treated cases of malaria are considered as reported due to private-sector treatment seeking. Based on the 2018-2019 MIS [29], about 44% of treatments were seen at private hospitals or clinics, with another 14% seeking treatment at private shops or pharmacies; an estimated 43% of treatments were seen in the public sector and reported. As such, for the target calibration year of 2023, there were an estimated 34.0 million clinical cases of malaria in the model, with 29.9 million treated, and 12.9 million reported in the public sector. While the model’s projection of total reported cases is low compared to the 14.2 to 16.0 million public sector clinical cases reported in Uganda in 2023 [30], the model’s prevalence projection is within the margin of error of the MIS district PfPR_0-59_ values from the simulation.

### 2.4 Model calibration to current kelch13 circulation

A total of 83 *pfkelch13* allele frequencies, for alleles 469Y and 675V, were compiled from public literature [18,31–34] covering the period from 2015 to 2022 in 15 districts in Uganda (Supp Table S3). To calibrate the model to these field data, simulations were run from 2004 to 2025, using the above calibrated prevalence and incidence values. Seven new model initialization parameters were added to reflect the following circumstances of 469Y and 675V emergence in the past:

- The district where 469Y first appeared
- The district where 675V first appeared
- The date of 469Y appearance
- The date of 675V appearance
- The initial allele frequency of 469Y (at appearance)
- The initial allele frequency of 675V (at appearance)
- The time at which mutation was activated in the model (after the model begins running in 2004, mutation is deactivated at least until equilibrium is reached)

Note that the term ‘appearance’ is intentionally vague, as we have no way of knowing when the first mutation occurred. In the simulation, we simply assume that one group of mutated parasites appears in a particular district at a particular time, with initial allele frequency *x* (which is fit below).

A total of 100 candidate parameter combinations were selected using Latin hypercube sampling to explore the relationships between the initial conditions and resulting allele frequencies during 2015-2022. For each parameter set, 30 model runs were executed with slight jittering to better capture the statistical variability between inputs and outputs, resulting in 3000 simulation runs. A machine learning approach, employing a traditional neural network built in PyTorch [35] was used to map the relationship between model inputs and the allele frequencies observed in 15 districts in Uganda. Two calibration approaches were applied:

1. **Data-to-Parameters mapping**: with this approach, the final calibrated neural network maps allele frequency data to the seven parameters listed above and the true allele frequency data are used to infer model initialization parameters.
2. **Parameters-to-Data mapping**: the final calibrated neural network, similar to a ‘model emulation’ tool [36], is used to produce model outputs (allele frequencies) from input or initialization parameters (i.e. the seven parameters listed above). For this final calibrated neural network, we use an objective-function based search method to produce outputs close to the observed real-world data.

The “D-to-P” approach results in the development of an NN_D-to-P_ model that, once calibrated, takes allele frequency data as input and generates the corresponding model initialization parameters as output. One has to verify in this case that the model, when initialized with these parameters, produces the correct allele frequency output. In contrast, the P-to-D approach constructs an NN_P-to-D_ model that emulates the simulation process, allowing the neural network to be calibrated using an objective function centered on the true data (a feature not achievable with the NN_D-to-P_ approach). Building, calibrating, and training the NN_P-to-D_ model introduces an additional step where input parameter values are selected based on the best fit to the data. This fit was determined using three distinct objective functions, with absolute mean deviation between allele frequencies (83 pairs) serving as the primary objective function. A small penalization factor (*c* ≤ 1.0) was applied to real-world allele frequencies that were lower than the model-predicted values, reflecting the likelihood that late arrival of an allele in a district is more probable than early arrival, due to the asymmetric variance associated with any waiting-time process.

The NN_P-to-D_ model was optimized using the Optuna Python library [37]. The process involved iterations from five random starting conditions (using Latin hypercube sampling boundaries), with comparisons of outputs for penalization factors *c* = 0.1, 0.5, and 1.0; this revealed no qualitative differences in the model fits as observed visually. The *c* = 0.5 penalty was chosen for the final calibrated NN_P-to-D_ model, and the inferred initialization parameters were: the 469Y allele was introduced on June 3, 2006, in Kumi district, with an initial frequency of 0.0534, while the 675V allele was introduced on December 10, 2006, in Soroti district, at a frequency of 0.000451. Mutation in the model was activated on October 27, 2012. This model was run and visualized to ensure that the projected allele frequencies (Figure 2) and prevalence equilibrium matched real-world conditions between 2013 and 2022. Note that the initial appearance times are not meant to correspond to the true times that these allele frequencies were reached, as model simulations typically show an initial decrease after initial (artificial) appearance, followed by a standard expected sustained increase due to the action of natural selection. Mutation rates were calibrated as blood-stage asexual-parasite mutation events as described in previous modeling exercises [38,39].

### 2.5 Treatment Strategies

Model runs were burned in for 10 years to achieve equilibrium (2004-2014) and then run under the treatment coverage settings in Table 1 (for another ten years, 2014-2025), with 43.0% of patients receiving AL in the public sector, 41.2% receiving AL in the private sector, and the remainder receiving sulfadoxine-pyrimethamine (2.1%), chloroquine (1.9%), artesunate monotherapy (4.8%), quinine (0.8%), or amodiaquine monotherapy (6.2%). The post burn-in calibration period of the model was run until July 1 2025, at which point the prevalence and incidence were checked against available data as shown in Figures 1C and 1D. On July 1 2025, the model either (1) continued to be run under primarily AL use as a status quo simulation of using AL as first-line therapy for uncomplicated falciparum malaria, or (2) was run under an alternate treatment strategy implemented in the public sector only. Model runs completed on Jan 1 2032 and comparisons between treatment strategies and the status quo were done over the six-year period from July 1 2025 to July 1 2031.

**FIGURE 1.**
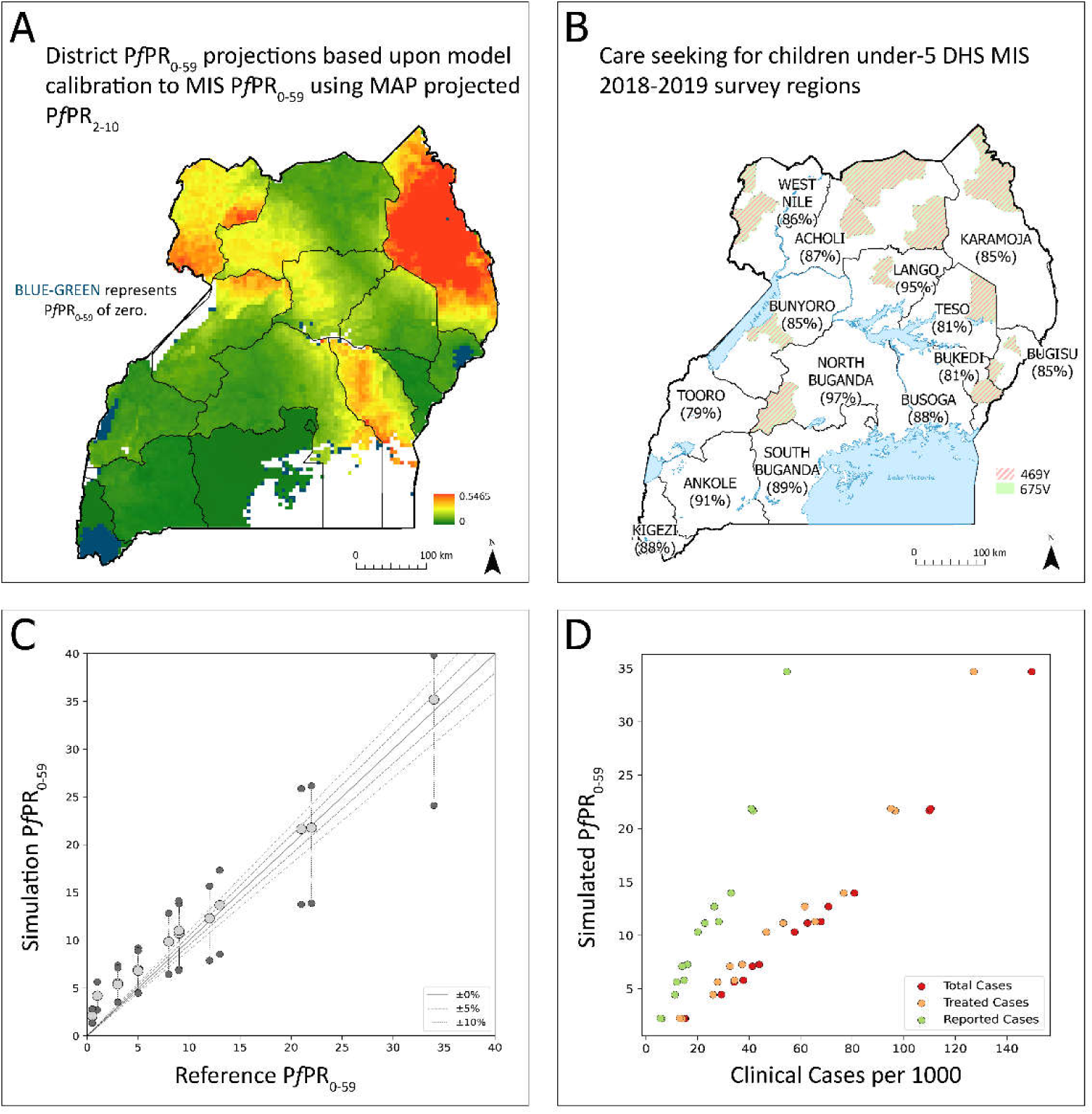
**(A)** Final PfPR_0-59_ calibration derived from Malaria Atlas Project estimates for PfPR_2-10_ in 2017, and later adjusted to Malaria Indicator Survey PfPR_0-59_ values from 2018-2019 [29] (page 55). Map prepared by authors using simulation outputs, Malaria Indicator Survey 2018 DHS Regions Boundaries [21], and Uganda Boundaries [20]. **(B)** Treatment coverage for children under five, according to 2018-2019 MIS. Shaded districts show districts where *pfkelch13* alleles 469Y or 675V have been identified. **(C)** Comparison of model-simulated prevalence in MIS districts to MIS data. The gray dot shows the annual mean prevalence in 0-5 year-olds while the dark gray dots at top and bottom indicate the annual upper and lower bounds. (**D**) By MIS district, the number of simulated malaria cases per year (red), the number of simulated malaria cases that are treated (yellow), and the number of treated cases that are likely to be reported to the public health system (green) based on treatment seeking choices made between private and public sector. Reported clinical cases are plotted as annual incidence per 1000 individuals (x-axis) and plotted against the simulated PfPR_0-59_ (y-axis).

**Figure 2:**
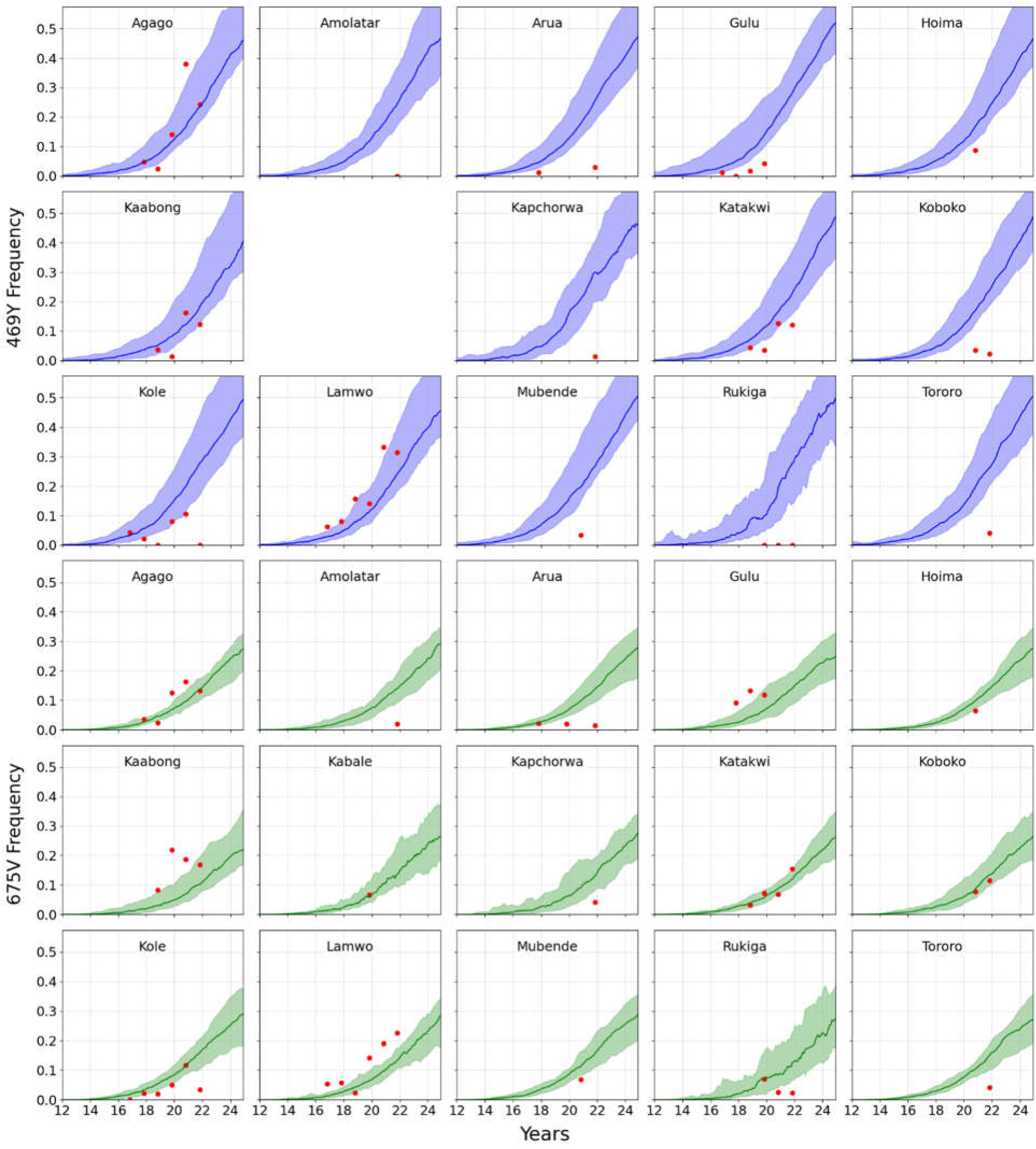
Calibration of *pfkelch13* 469Y and 675V allele frequencies (NN_P-to-D_, *c* = 0.5) in 15 districts in Uganda. Each panel shows the simulated 469Y allele frequency (blue line) or 675V allele frequency over time, in one district, with the shaded area representing the interquartile range of the simulation outputs. The red dots correspond to the observed data collected between 2015 and 2022. The model performs well in districts such as Kaabong, Katakwi, and Lamwo, where the predicted frequencies closely follow the observed data. In contrast, early predictions of allele emergence are evident in districts like Kole, Hoima, and Tororo, where the model anticipates a faster rise in frequency compared to the true data. Note that the one sample in Kagale district [40] was sampled at the same health facility as samples from Rukiga district; around the time of sampling Kabale district was split into two districts (Kabale and the new district of Rukiga).

Two of the evaluated treatment strategies were first-line therapy switches to either ASAQ or DHA-PPQ. Cycling strategies were evaluated with pre-planned rotation schedules. All possible cycling approaches with two or three ACTs and with 1-year/2-year/3-year rotation periods were considered, for a total of 30 strategies. MFT strategies were implemented as multiple therapies deployed in the same 5km-by-5km pixel, with the model using a simple random draw – e.g., under status quo, patients were given AL with 84.2% probability, artesunate monotherapy with 4.8% probability, etc. – to assign a therapy to a particular patient with symptomatic confirmed malaria. All MFT strategies with 50/50 and 75/25 use percentages of two ACTs were considered, along with a 33/33/33 strategy where all three ACTs were used in equal proportion (a total of ten strategies). Ten strategies were run with the triple ACT artemether-lumefantrine-amodiaquine (ALAQ). These strategies included immediate deployment, deployment delayed by four years, and deployment delayed by four years with an interim cycling strategy used before ALAQ deployment. A total of N=32 simulation runs were performed for each strategy. No geographically stratified approaches were considered in this analysis.

## 3 Results

The Uganda-calibrated individual-based simulation was run to July 1, 2025, achieving a nationwide parasite prevalence in 2-10 year-olds (PfPR_2-10_) of 17.5% (90% range: 17.4% - 17.6%) and a nationwide reported incidence of 263 malaria cases per 1000 individuals per year. This corresponds to 12.9 million (90% range: 12.83M - 12.96M) malaria cases reported to the public sector in 2023. It is assumed that 84% of patients received AL for uncomplicated falciparum malaria and that the remaining 16% received one of sulfadoxine-pyrimethamine, chloroquine, artesunate monotherapy, quinine, or amodiaquine. All results for treatment failure counts, treatment failure percentage, and allele frequencies of *pfkelch13* mutations are presented from July 1 2025 to July 1 2031.

### 3.1 Status Quo

The continuation of artemether-lumefantrine (AL) as first-line malaria treatment in Uganda is projected to result in increases in malaria burden and substantial increases in treatment failures over the next six years. The projection from our Uganda-calibrated malaria simulation model is that from 2025 to 2031, taking the median across all simulation runs, Uganda will experience an annual average of 32.8 million malaria cases, with an annual average of 9.9 million treatment failures or a monthly average of 820,898 treatment failures; the monthly average over six years is used here as our quantity for comparison. This assumes no changes in the amount and effectiveness of malaria control activity, only changes in the levels of circulating drug-resistant genotypes. For the first year of the evaluation period (July 1 2025-July 1 2026) the model projects a median of 31.2 million malaria cases, with 26.2% treatment failure at the population level across all treatments taken, and in the final year of model evaluation (July 1 2030-July 1 2031) the model projects a median 34.5 million malaria cases with 33.1% population-level treatment failure. The monthly treatment failure count (Figure 3) increases from 684,687 during the first year of implementation to 952,335 over the final year of implementation, corresponding to an annual 6.8% increase in total treatment failures over this period. Note that these treatment failure percentages are being reported for an entire population, across public and private markets and across all drugs taken, and they are not comparable to treatment failure rates inferred from therapeutic efficacy studies. The primary reason for the increase in treatment failure percentage is the continued use of a single ACT (artemether-lumefantrine) which is predicted to rapidly select for currently circulating kelch variants and any alleles associated with decreased susceptibility to the partner drug lumefantrine.

**Figure 3:**
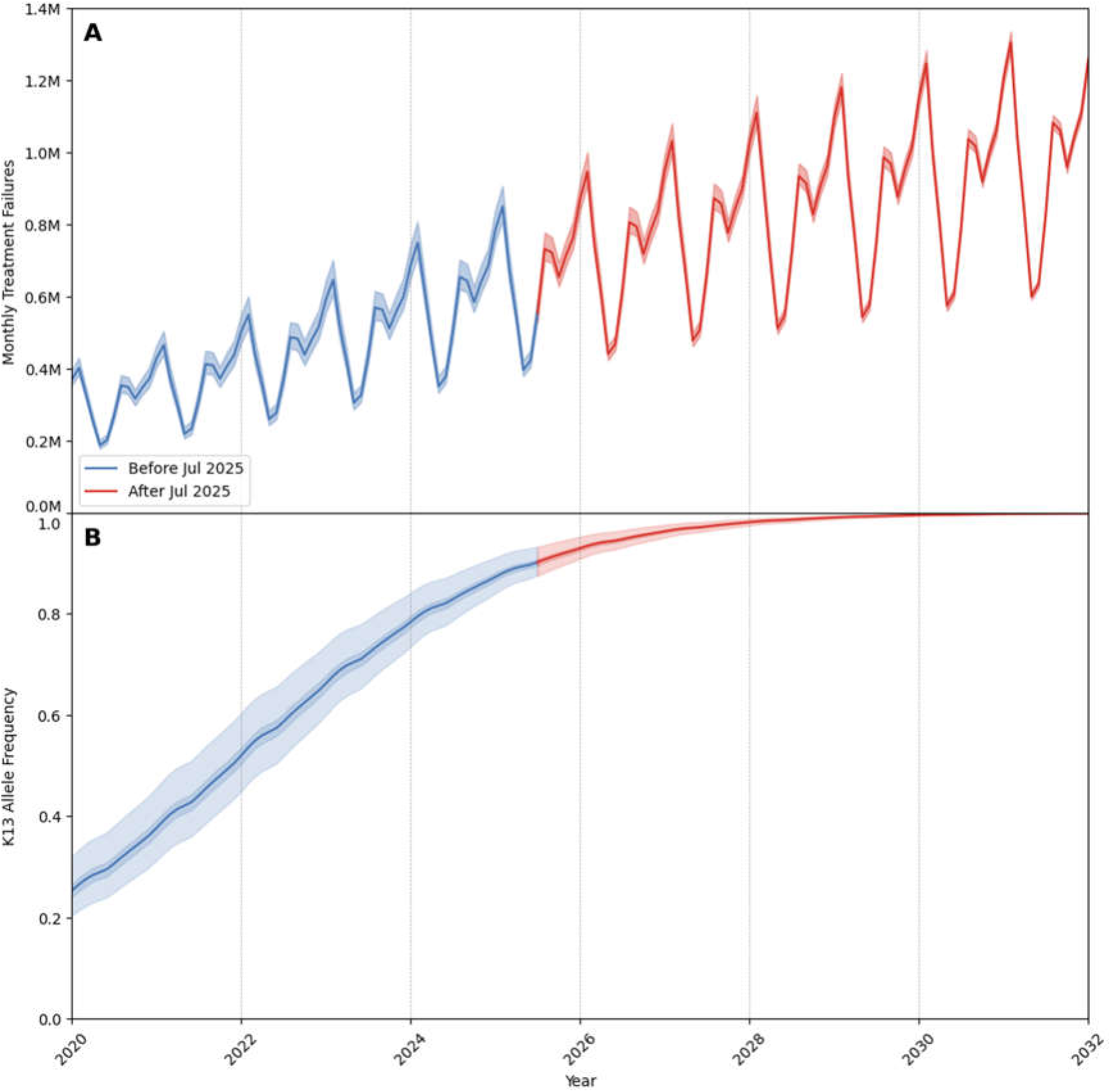
Trends in malaria treatment failures and *pfkelch13* allele frequency (2020–2032) under the status quo strategy in Uganda of continued artemether lumefantrine use. Blue lines represent model calibration prior to July 2025, while red lines show model projections after July 2025. Panel A shows a steady increase in monthly treatment failures over time, surpassing one million by 2030 as resistance to both artemisinin and lumefantrine become widespread. Panel B shows the combined *pfkelch13* allele frequency (sum of frequencies for two alleles) which is estimated to reach 0.90 by July 2025. Shaded regions denote the 50% and 90% ranges across simulations illustrating variability in resistance dynamics.

The rapid increase in artemisinin partial resistance can be seen in the projected sharp rise in the *pfkelch13* allele frequency (Figure 3B). In July 2025, the combined allele frequency of *pfkelch13* alleles 469Y and 675V in Uganda is already estimated to be at a median of 0.91 (90% range: 0.88 - 0.94) suggesting that a large portion of the parasite population carries one of these two mutations. Note that for 131 out of Uganda’s 146 districts, there are no data on *pfkelch13* allele frequencies, and the model’s migration mechanism causes these two kelch variants to spread evenly to nearly all districts over the >10 years since their emergence (it is not known if this accurately reflects the true migration process of these variants). From 2025 to 2031, under the status quo, the combined allele frequency of these two kelch variants is projected to increase to 1.0 (90% range: 1.0 - 1.0) simply because they are under strong selection; all population-genetic and evolutionary models would project fixation of these phenotypes under these circumstances (see limitations section of Discussion).

### 3.2 Switches of first-line therapy to ASAQ or DHA-PPQ

The first strategies to consider in the face of rising drug-resistance are first-line switches to a different ACT, either to artesunate-amodiaquine (ASAQ) or dihydroartemisinin-piperaquine (DHA-PPQ). These two strategies show distinct impacts on the projected number of malaria cases, treatment failures, and the spread of artemisinin resistance. Both options reflect potential improvements over the status quo, though they come with trade-offs. Under a switch to ASAQ, the total annual average of malaria cases from 2025 to 2031 is projected to be 32.8 million, a 0.3% reduction compared to cases expected under AL. However, ASAQ provides a significant reduction in the annual number of treatment failures, estimated at 6.3 million compared to 9.9 million under AL (a 36.4% reduction). In the first year of the intervention, monthly treatment failures are projected to be 468,881 (90% range: 451,836 – 482,796), rising to 582,052 (90% range: 568,016 – 595,055) in the final year (July 2030 to July 2031); see Figure 4A. For the entire period from 2025 to 2031, the average monthly treatment failures under ASAQ deployment are projected to be 522,456 (90% range: 506,818 – 535,748). The combined frequency of the 469Y and 675V alleles is estimated to be 0.91 (90% range: 0.88 - 0.94) in July 2025, and increasing to 0.99 (90% range: 0.99 - 1.0) by July 2031, as expected under any policy continuing to use ACTs (Figure 4B). While the spread of artemisinin partial resistance is projected to continue under ASAQ deployment, treatment failures will be less common, as the partner-drug resistance landscape will be favorable to amodiaquine treatment even in the presence of kelch13 mutations. This suggests that switching to ASAQ should provide a short-term and potentially medium-term solution for mitigating resistance, though it is unlikely to slow the progression of artemisinin-resistant genotypes.

**Figure 4.**
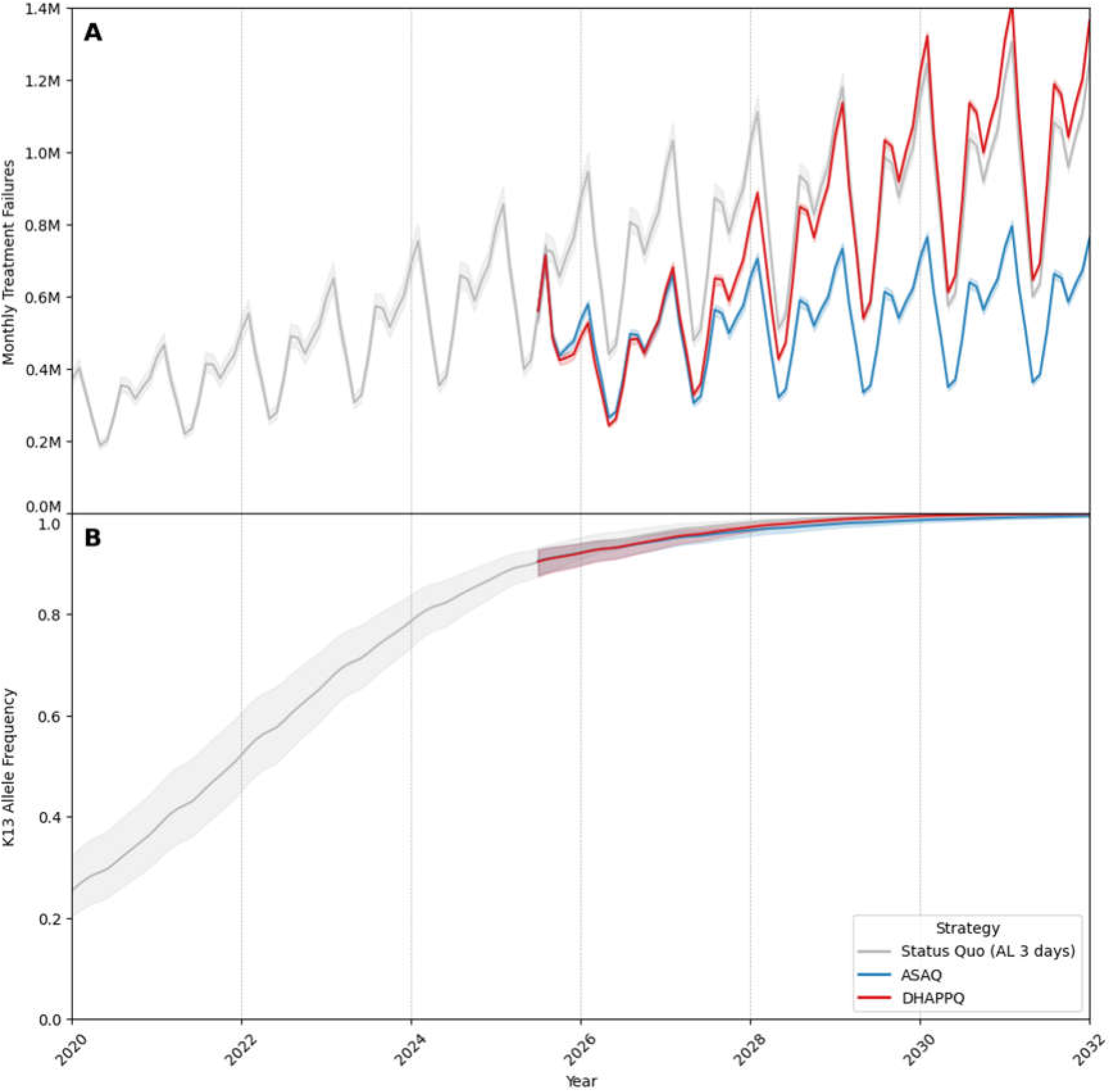
Monthly treatment failures and *pfkelch13* allele frequency under three treatment strategies (2020–2032). This figure compares the projected monthly treatment failures (Panel A) and *pfkelch13* allele frequency (Panel B) under three malaria treatment strategies in Uganda: the status quo (continued use of artemether-lumefantrine, gray), switching to artesunate-amodiaquine (ASAQ, blue), and switching to dihydroartemisinin-piperaquine (DHA-PPQ, red) in July 2025. Panel A shows that after July 2025, DHA-PPQ exhibits the steepest temporary reduction, followed by a sharp rise after 2028, surpassing the status quo treatment failure count by 2030. ASAQ maintains lower treatment failures compared to the status quo throughout the simulation period. Panel B shows the progression of combined *pfkelch13* allele frequency (sum of frequencies for two alleles) over the same period. Post-switch, resistance accelerates at a similar rate under DHA-PPQ, ASAQ, and the status quo, with all three treatment strategies showing comparable trajectories and reaching fixation by 2030. Shaded regions represent the 90% uncertainty range, reflecting variability across simulations.

In contrast, switching to DHA-PPQ as first-line therapy results in a lower overall malaria burden but with substantial additional risk in terms of treatment failure increases. From 2025 to 2031, the annual number of malaria cases under DHA-PPQ is projected to be 31.5 million, representing a reduction of 3.9% compared to AL. However, the total treatment failures are projected to be 31.5 million over the same six-year period. Monthly treatment failures are projected to be 443,319 (90% range: 429,965 – 457,492) in year one of the intervention and to rise substantially to 1,036,535 (90% range: 1,021,732 – 1,046,013) by 2030-2031. For the 2025 to 2031 period, the average monthly treatment failure count for DHA-PPQ deployment is 728,884 (90% range: 715,408 – 738,795) showing a moderate decrease (11.2% reduction) when compared to status quo AL use. Kelch variants are again projected to reach fixation by 2031 (Figure 4B). Although DHA-PPQ may initially reduce malaria cases and prevalence more effectively than ASAQ or AL, the projected rapid evolution of piperaquine resistance diminishes the long-term utility of DHA-PPQ. It is important to state that, as in our previous analysis for Rwanda [11], the piperaquine-resistant genotypes in our model are parameterized via known PPQ-resistant phenotypes in SE Asia whose failure rates reached 58% [41]. It is not known if these same high-failure PPQ-resistant *P. falciparum* phenotypes will evolve in Africa, therefore there is substantial uncertainty in these treatment faure projections under DHA-PPQ deployment.

### 3.3 2-year cycling approaches

Switching from the status quo to a 2-year cycling strategy in 2025 is projected to substantially reduce malaria treatment failures for most cycling sequences and choices of ACT. Figure 5 shows a summary of monthly treatment failure counts averaged over the six years from 2025 to 2031 for all twelve possible ways to cycle AL, ASAQ, and DHA-PPQ. A cycling approach like this can use either two ACTs or three ACTs. The bottom half of Figure 5 shows that including all three ACTs in the cycling regimen results in similar outcomes, regardless of which therapies are used first, second, and third. When two ACTs are used, the projected number of treatment failures depends strongly on which two therapies are chosen. Note that these cycling policies apply only to the antimalarials prescribed in the public sector (43% of all treatments), while AL continues to be used in the private sector for the period of evaluation. The pace of *pfkelch13* evolution is similar for all twelve cycling options.

**Figure 5.**
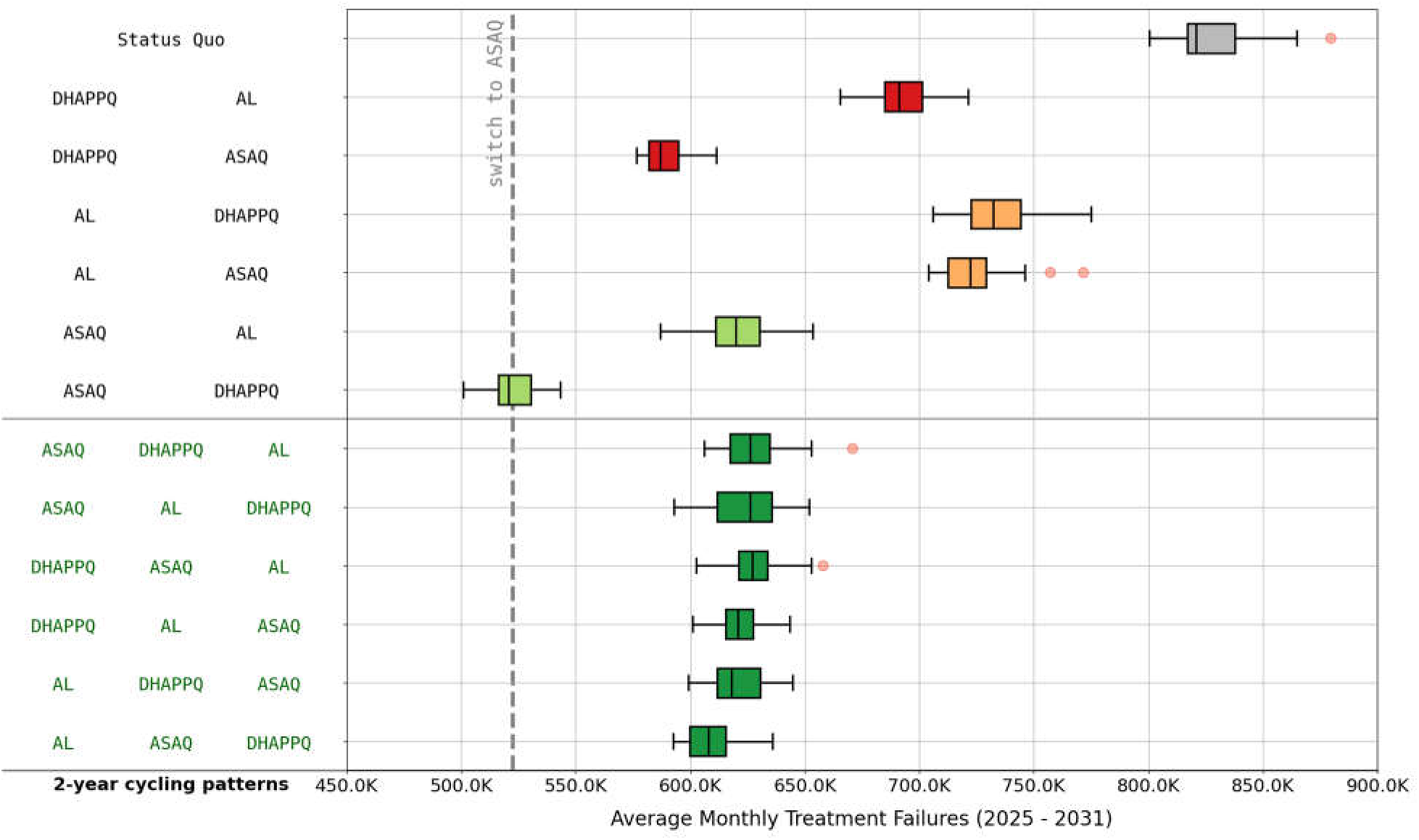
Average Monthly Treatment Failures (2025 - 2031) under Different 2-Year Cycling Strategies. This box plot illustrates the projected average monthly treatment failures for various 2-year cycling regimens using artemether-lumefantrine (AL), artesunate-amodiaquine (ASAQ), and dihydroartemisinin-piperaquine (DHA-PPQ). The vertical dashed line represents the median monthly treatment failures for a single-drug switch to ASAQ after July 2025. Light green boxes correspond to two-drug cycles starting with ASAQ, red boxes represent two-drug cycles initiated with DHA-PPQ, orange boxes indicate two-drug cycles starting with AL, and dark green boxes represent three-drug cycling strategies. While three-therapy cycling strategies achieve low treatment failure counts, none of these outperforms a two-therapy cycle of ASAQ followed by DHA-PPQ.

**Figure 6.**
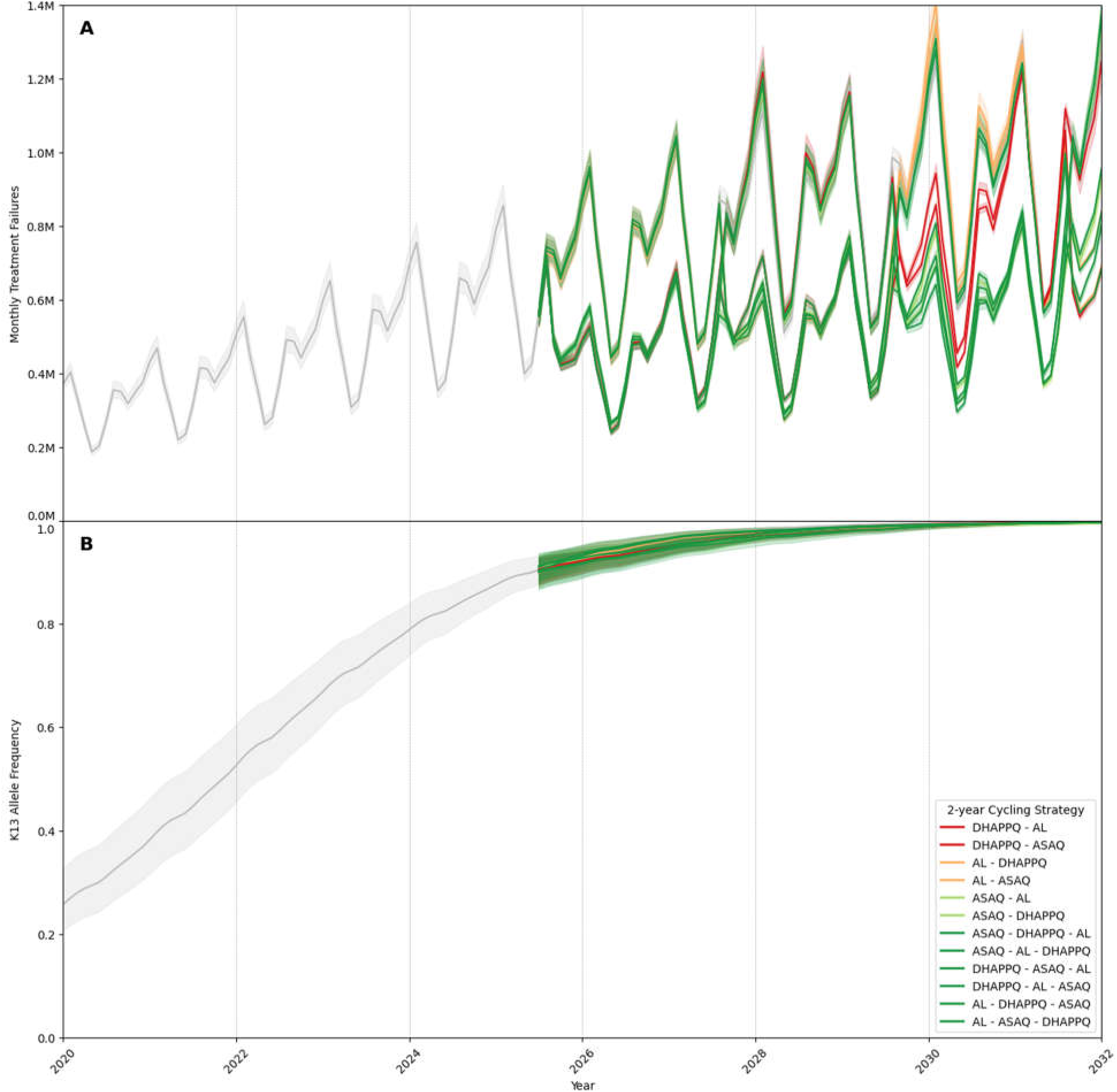
Monthly treatment failures and *pfkelch13* allele frequency under different 2-year cycling strategies (2020–2031). This figure compares the projected monthly treatment failures (Panel A) and *pfkelch13* allele frequency trajectories (Panel B) under various 2-year cycling strategies involving artemether-lumefantrine (AL), artesunate-amodiaquine (ASAQ), and dihydroartemisinin-piperaquine (DHA-PPQ). Dark green lines represent cycling strategies involving all three therapies (AL, ASAQ, DHA-PPQ). Light green lines correspond to two-therapy cycling strategies starting with ASAQ. Orange lines represent two-therapy cycling strategies starting with AL. Red lines show two-therapy cycling strategies starting with DHA-PPQ. Shaded regions in both panels represent the 90% uncertainty range across simulations.

The optimal strategy for a two-year cycling approach is a two-therapy cycle of ASAQ and DHA-PPQ, with ASAQ deployed first. Under this strategy, the projected average monthly treatment failure count for 2025–2031 is 520,568 (90% range: 508,997 – 537,678), representing a 36.6% reduction compared to the status quo. The second-best strategy starts with DHA-PPQ and cycles to ASAQ; this yields a monthly treatment failure count of 587,119 (90% range: 577,077 – 605,825) which is a 28.5% reduction compared to the status quo. Note that in both of these strategies, AL is used in the private market, meaning that at any point in time the on-the-ground treatment usage is either a well-mixed MFT policy with DHA-PPQ and AL or a well-mixed MFT policy with ASAQ and AL. This is likely one of the reasons that these two approaches are associated with the lowest number of projected treatment failures. Among two-therapy strategies, regimens incorporating AL first are projected to have the highest monthly treatment failure counts, ranging from 723,369 to 732,470, again largely attributable to the substantial use of AL in the private market, resulting in four out of six years where AL is used for ∼84% of treatments.

When all three ACTs are included in the cycling strategy, the projected average monthly treatment failure count falls in a narrow range from 607,906 to 627,094, showing a 24% to 26% improvement over status quo of AL use. However, none of these approaches is able to outperform the optimal cycling strategy of ASAQ first and DHA-PPQ second, which takes advantage of (1) the opposing resistance phenotypes seen at the *pfcrt* and *pfmdr1* loci, which make simultaneous drops in efficacy to amodiaquine and lumefantrine difficult to achieve, and (2) the constancy of AL use in the private market, which allows for a private-public MFT deployment when a non-AL ACT is used in the public sector.

Finally, as can be seen plainly in Figure 5, an ASAQ-DHAPPQ 2-year cycling approach is only as good as a straightforward switch to six years of ASAQ use. This is because the current set of circulating genotypes in Uganda (*pfcrt* K76, *pfmdr1* N86) will be acted on most efficaciously by ASAQ, with a 2022-2023 therapeutic efficacy study in eastern Uganda showing 100% efficacy of ASAQ [42]. The reason for this is that genotypes with reduced sensitivity to lumefantrine have been selected for over the past 20 years (in most African contexts) and these genotypes – typically K76 in *pfcrt* and N86 in *pfmdr1* – are the ones on which ASAQ has the highest efficacy. This means that most cycling approaches for Uganda can be redesigned around using ASAQ for a longer period first – i.e. a transition period where ASAQ is used as first-line therapy – while operational plans are put into place to transition to a longer-term cycling approach among ACTs and potentially non-ACT therapies if they are made available.

The benefits of cycling approaches with 1-year or 3-year cycles are essentially dependent on how often AL is used during the six-year period from 2025 to 2031 (Supp Figures S2 and S3). When AL is omitted from the cycling regimen, these approaches also result in a projected average of 540,000 to 550,000 monthly treatment failures, slightly less optimal than the 2-year cycling approach with ASAQ followed by DHA-PPQ.

### 3.4 Multiple First-line Therapies

As a standard comparison to drug cycling, we evaluate the deployment of two or three ACTs simultaneously, also referred to as multiple first-line therapies of MFT [9,43]. Note that in these modeling assessments, it is assumed that patients randomly receive one of several ACTs, and no mechanisms of distribution by age, health facility, or region are used. The standard uniform MFT approach, where each third of the population is treated with one of three ACTs, results in 624,462 monthly treatment failures (90% range: 606,832 – 644,643), which is a moderate improvement over the status quo (23.9% reduction) but remains higher than the most effective MFT strategies. As in previous analyses [19], the perfectly uniform MFT approach is not always best among all considered strategies, as current partner-drug landscape and future resistance risk are not equal among the three ACTs.

Among MFT options, strategies with lower AL use consistently result in fewer treatment failures compared to those with higher AL use. Figure 7 ranks these strategies by monthly treatment failure count, clearly showing that MFT approaches where 75% of patients receive AL for six years (in red) lead to the highest numbers of treatment failure rates, averaging 750,000 per month. While this represents an 8.6% improvement over the status quo, it remains substantially worse than the best-performing MFT strategies. MFT strategies where 25% to 50% of patients receive AL for six years (in orange) produce monthly treatment failure counts ranging from 600,000 to 700,000 – this is still suboptimal when compared to certain treatment strategies projected to have 520,000 monthly treatment failures. These results emphasize the importance of reducing public-sector AL use while AL is still being widely used in the private sector.

**Figure 7.**
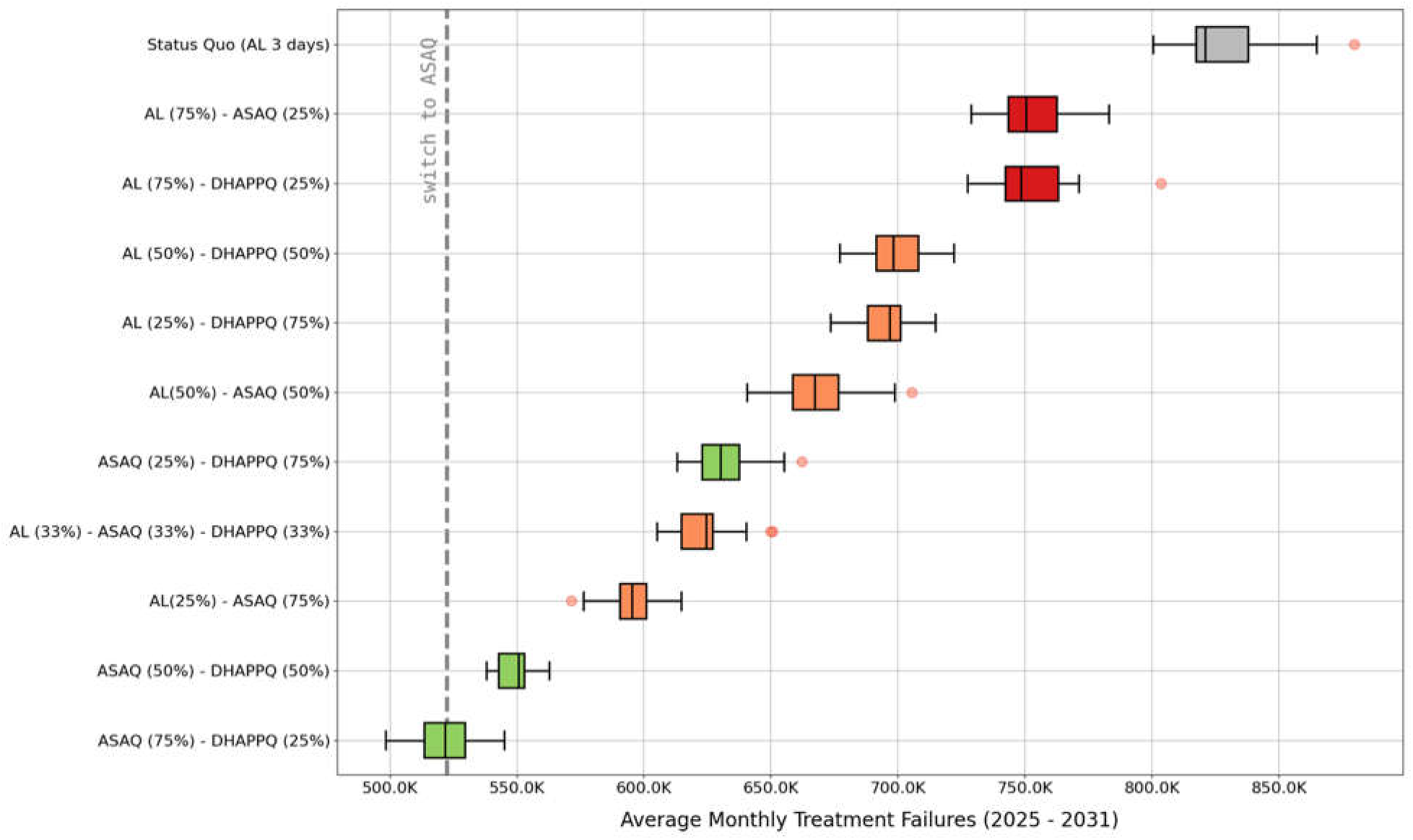
Average monthly treatment failures (2025-2031) across different MFT strategies. Box plots show the average monthly treatment failure counts from 2025 to 2031 under various multiple first-line therapies (MFT) strategies with different proportions of artemether-lumefantrine (AL), artesunate-amodiaquine (ASAQ), and dihydroartemisinin-piperaquine (DHA-PPQ) deployed in the public sector. The status quo corresponds to continued use of AL and has the highest median monthly treatment failure count at 820,898. MFT strategies are color-coded based on AL usage: green represents strategies with no AL; orange shows strategies where AL represents 25% to 50% of treatments; red shows strategies where AL is used for 75% of malaria cases. The vertical dashed grey line represents the median monthly treatment failures observed for a first-line switch to ASAQ. The MFT strategies with the lowest average monthly treatment failures is ASAQ (75%) - DHA-PPQ (25%), achieving a median monthly treatment failure count of 521,649, a 36.5% reduction compared to the status quo.

**Figure 8.**
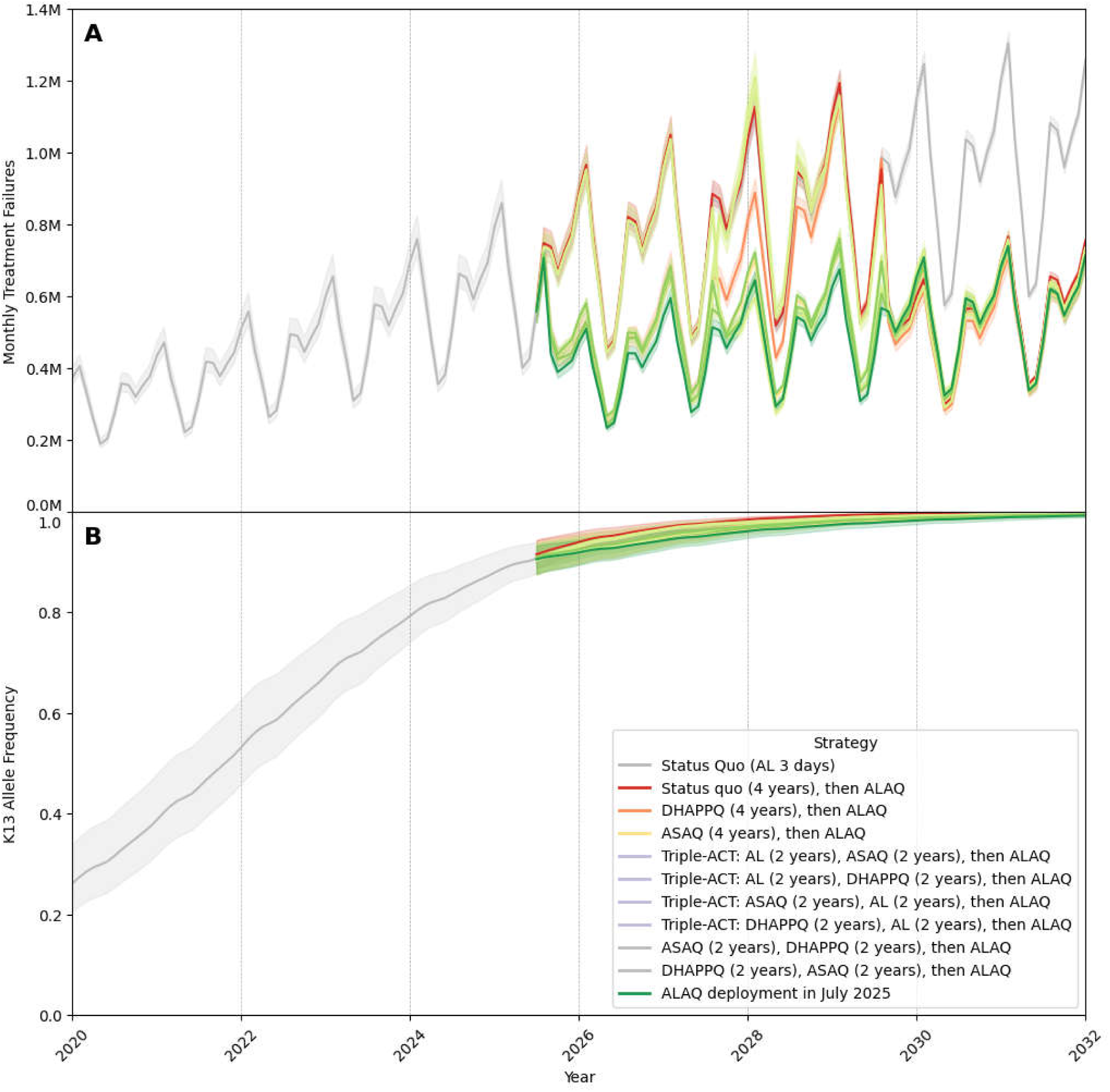
Monthly treatment failures and *pfkelch13* allele frequency under different triple ACT (ALAQ) deployment scenarios through 2031. This figure compares the projected monthly treatment failures (Panel A) and *pfkelch13* allele frequency trajectories (Panel B) under various triple ACT (ALAQ) deployment strategies. The gray line represents the status quo, while the dark green line corresponds to immediate deployment of ALAQ, which achieves the best outcomes by keeping the average monthly treatment failure count below 500,000 and somewhat slowing the fixation kelch mutations. The dark grey lines show 2-year cycling approaches using ASAQ and DHA-PPQ before switching to ALAQ. The orange line (DHA-PPQ), yellow line (ASAQ), and red line (AL) represent scenarios where a single ACT is used for four years before transitioning to ALAQ. In Panel B, combined *pfkelch13* allele frequency is shown. Shaded regions in both panels represent the 90% ranges of simulations.

The optimal MFT choice in Figure 7 combines ASAQ (75% use in the population) and DHA-PPQ (25% use), taking advantage of the beneficial partner-drug resistance landscape to amodiaquine and a relatively low level of PPQ use to weaken the selective pressure on piperaquine-resistance evolution. This approach is projected to generate an average of 521,649 treatment failures (90% range: 505,669 – 538,608) per month, corresponding to a 36.5% reduction compared to status quo. Thus, the best MFT strategy has a nearly identical treatment failure outcome as (1) the best cycling strategy and (2) a switch to first-line ASAQ use, again underscoring the utility of using ASAQ as a transitional therapy for the second half of the 2020s.

### 3.5 Options for triple ACT deployment

Finally, the largest reduction in near-term and medium-term treatment failure counts is projected to result from deployment of the triple therapy artemether-lumefantrine-amodiaquine (ALAQ). This is consistent with projections from a past multi-model comparison [44], our analysis of ALAQ deployment in Rwanda [11], and evaluation of ALAQ deployment with a new more complex 25-locus individual-based model of *P. falciparum* evolution [45]. Switching to a triple artemisinin-based combination therapy (TACT) offers a promising avenue for reducing malaria treatment failures and slowing the spread of resistance. However, the timing of the transition — whether immediate or delayed — affects this strategy’s efficacy. While immediate implementation of ALAQ is the most effective strategy [44], the time required for regulatory approval, manufacturing, and establishment of supply chains will necessitate interim approaches. Here, we also simulate a four-year delay before ALAQ is approved as a first-line therapy with alternate regimens used during the interim period.

An immediate switch to ALAQ yields the fewest treatment failures, with a monthly average of 477,929 (90% range: 463,464 – 499,508) which is 41.8% lower than the status quo forecast of 820,898. If ALAQ is not available until July 2029, the best interim approach is to switch to ASAQ for four years or implement a 2-year cycle of ASAQ followed by DHA-PPQ before transitioning to ALAQ. These strategies effectively balance resistance management and treatment efficacy, with projected monthly treatment failures averaging 503,067 (90% range: 493,290 – 519,556) under the four-year ASAQ approach (a 38.7% compared to status quo) and around 506,000-508,000 when cycling ASAQ and DHA-PPQ.

In contrast, continuing AL use for four years before introducing ALAQ is the worst option among the evaluated strategies, with monthly treatment failures reaching 706,494 (90% range: 687,993 – 736,851), still a 13.9% reduction compared to no TACT switch. While all TACT deployment options outperform the status quo, strategies that include 2-year cycling with AL (lavender box plots in Figure 9) before switching to ALAQ lead to higher treatment failures and delay the full benefits of ALAQ, reinforcing the importance of minimizing AL use in transition plans.

**Figure 9.**
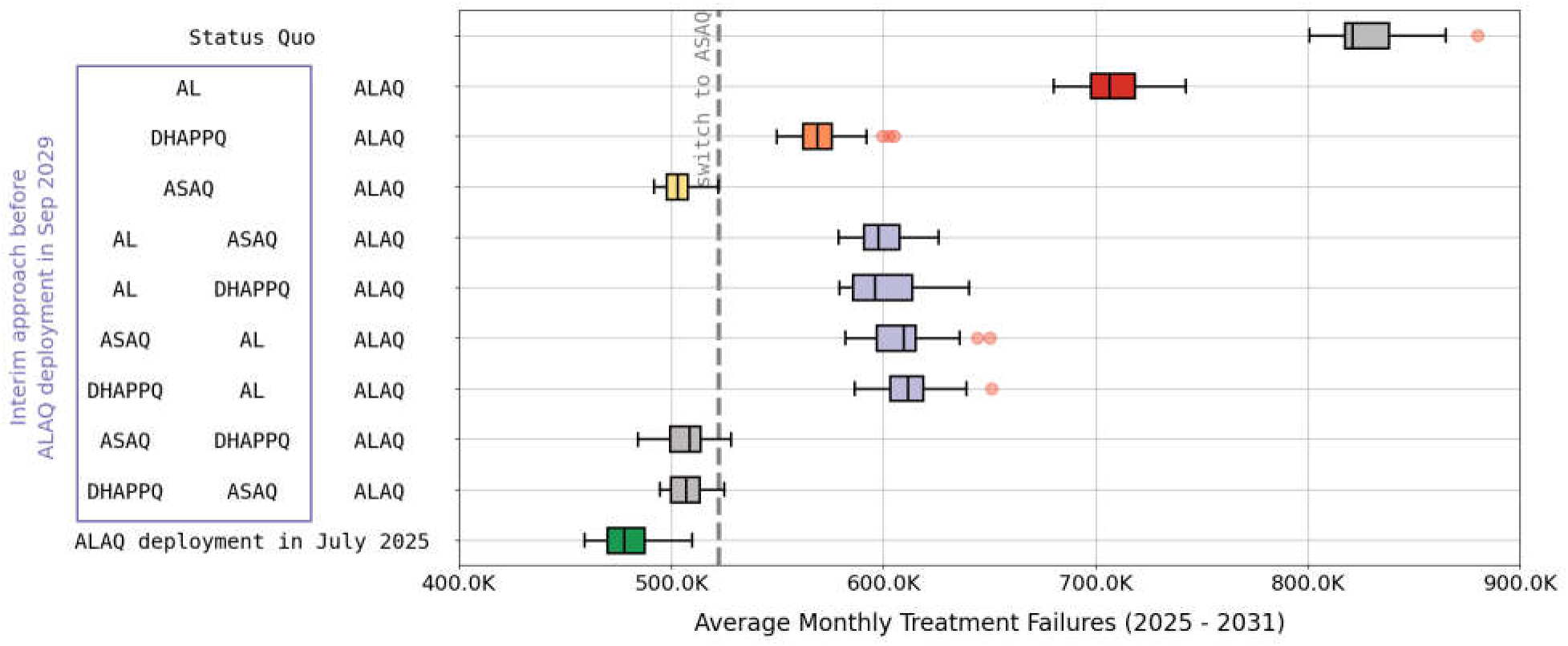
Average monthly treatment failures (2025-2031) under different triple ACT (ALAQ) deployment scenarios. Box plots show the average monthly treatment failure counts for the period 2025-2031 for various ALAQ deployment scenarios. The vertical dashed grey line represents the median monthly treatment failures observed when ASAQ is used as first-line therapy after July 2025. If ALAQ is not available until July 2029, the best interim approach is to switch to ASAQ for four years or implement a 2-year cycle of ASAQ and DHA-PPQ before transitioning to ALAQ. These strategies effectively balance resistance management and treatment efficacy, with projected monthly treatment failures averaging ranging from 503,000 to 508,000. These represent substantial reductions in treatment failure counts compared to the status quo. In contrast, continuing AL use for four years before introducing ALAQ results in the worst outcomes, with monthly treatment failures reaching 706,494 (90% range: 687,993 – 736,851). This comparison highlights the effectiveness of an immediate switch to ALAQ and underscores the advantage of minimizing AL use as an interim strategy when ALAQ deployment is delayed.

## 4 Discussion

As of 2023, among all countries in Africa, Uganda had the highest allele frequencies of *pfkelch13* mutations confirmed to be associated with artemisinin partial resistance. Five unique kelch13 variants had spread to at least 15 districts in Uganda by late 2022. There has been some evidence of ∼10% treatment failure rates to AL over the past ten years [46,47] and recent evidence of 15.4% and 17.1% PCR-corrected AL treatment failure in two districts in Uganda [42,48]; surprisingly, at each of these two sites the allele frequencies of *pfkelch13* mutations that mediate artemisinin partial resistance were low. The Ugandan National Malaria Elimination Division (NMED) has recognized the urgency of this situation and in 2023 began putting plans into place to mitigate the effects of spreading partial artemisinin resistance. The majority of strategies designed to counter the spread of resistance focus on altering the deployment of first-line antimalarial therapies in order to maximize current treatment efficacy against circulating *P. falciparum* genotypes and slow down the spread of mutations associated with partner-drug resistance. This report describes a first-phase evaluation of common treatment strategies available to the Uganda NMED in the second half of this decade.

Scenario evaluation results from our mathematical modeling analysis project that (1) continued use of AL as first-line therapy will result in an annual ∼7% increase in treatment failures over the next six years; (2) a switch to ASAQ as first-line therapy would reduce treatment failure counts by approximately 36.4% over this period – assuming prompt implementation and high levels of compliance; (3) cycling approaches and MFT approaches that are balanced toward higher ASAQ use and lower DHA-PPQ use will also lead to ∼36% reductions in projected treatment failure counts over a six-year period; and (4) immediate deployment of TACT would lower absolute treatment failure counts by ∼42% over the next six years. These standard national-level strategy designs have room for improvement in their adaptation to specific epidemiological contexts, adjustment for regional drug efficacy estimates, and enhancement by public health communication measures. If ASAQ deployment becomes a major part of Uganda’s partial artemisinin-resistance response in the 2020s, routine surveillance of AQ-resistance should be re-started.

The size of Uganda’s private antimalarial market has a substantial influence on the optimal choice of ACT deployment in the public sector. We assume that private-sector drug purchases cannot be influenced in the near-term by national-level treatment guideline changes, meaning that under all treatment scenarios we assume that AL will be used for 41% of all malaria cases and a non-ACT treatment will be used for another 16% of cases treated through private pharmacies or dispensaries. Our strategy comparisons indicate that we can take advantage of these fixed preferences in the private sector and recommend ASAQ or DHA-PPQ use in the public sector to achieve a locally mixed deployment of therapies with combining private-sector AL and public-sector ASAQ or DHA-PPQ (note that this approach would change if private-market preferences changed). The assumption in the modeling is that public clinics/dispensaries and private pharmacies co-exist in the same spaces and that there are no strong local preferences to seek treatment in a private versus a public facility. The modeling outcome is that different public-sector and private-sector treatment regimens will slow the increase in drug-resistance associated treatment failures because local use of two (or three) ACTs ensures that a patient has a 1-in-2 (or 2-in-3) chance of using an ACT whose partner-drug has high efficacy against recently selected partner-drug resistant genotypes. The general conclusion from this dynamic is that optimal strategies for the near-term and medium-term are those that recommend high or even 100% ASAQ use in the public sector. A reasonable and near-optimal long-term approach would be to use ASAQ as a transitional therapy until triple therapies or non-ACT combinations become available.

Considering changes to policy in Uganda, there is no guarantee that a geographically uniform national-level policy will be optimal among all available strategy choices, and there is likewise no guarantee that a regionally-targeted and context-specific design would have the high compliance and friction-free implementation needed to be as effective as projected in a mathematical model. Nevertheless, locally-targeted strategies must be considered, including specific designs for low-prevalence areas amenable to elimination efforts and increases in vector control, neither of which is considered in this analysis. In general, if it is known that either (a) the history of therapeutic use in a region or (b) current data on circulating genotypes in a region suggest that some ACTs are likely to have higher regional efficacy than others, a customized region-specific artemisinin-resistance response strategy should be considered. A second phase of treatment strategy design should evaluate combinations of locally-tailored approaches, cycling and MFT approaches, and deployment of novel therapeutics when they are made available.

A key tool for any resistance mitigation effort in the 2020s is deployment of artesunate-pyronaridine, an ACT that has the advantages of high efficacy [13,49] and no known resistance genotypes and the disadvantages of high price and difficult procurement. Pyronaridine does not appear to generate rapid or high-grade resistance in vitro. ASPY deployment is an ideal option in an MFT or cycling approach whose aim is to lower the resistance pressure on currently deployed partner drugs and also increase treatment efficacy of the therapies that are in use in the population. Introduction of ASPY would allow AL to be removed from use to a greater degree and would relieve the pressure on amodiaquine and piperaquine resistance evolution, both having occurred in the past, with large effects treatment efficacy. The challenge of increasing supply, easing procurement, and reducing the price of ASPY should be viewed by the global health community as an urgent priority for malaria management in the 2020s and beyond.

### Limitations

The major limitations in this analysis center on (1) our inability to precisely match genotypes to drug-resistance phenotypes, and (2) the lack of predictability around which individual mutations will arise first in certain contexts. An area of particular uncertainty is our limited knowledge on the complex evolutionary pathways that lead to piperaquine resistance and which of these pathways is the most likely to occur in African malaria settings. Four mutations in the *pfcrt* gene (T93S, H97Y, F145I, I218F) rose to high enough frequency in Cambodia that their effects on reduction or PPQ efficacy could be measured [50], but it is not known which of these or potentially other mutations with in vitro associations with PPQ-resistance [51–54] will be the first to emerge and be selected for in Uganda (or elsewhere in Africa). As stated in our previous analysis on this topic [11] the uncertain future of piperaquine resistance evolution in Africa is the major risk that needs to be managed as multiple ACTs are deployed in response to rising frequencies of kelch variants. This is not to say that DHA-PPQ should not be deployed as a component of artemisinin resistance response in Africa. On the contrary, DHA-PPQ deployment will likely play a critical role in any successful response by increasing near-term treatment efficacy. However, introduction and scale-up of DHA-PPQ should aim for moderate levels of DHA-PPQ usage at a population-level [9], and DHA-PPQ deployment should be accompanied by rapid-turnaround real-time molecular surveillance for known resistance markers.

Second, compliance and implementation are persistent challenges for any guideline change for infectious disease management. Several recent studies have aimed to collect qualitative information on compliance with treatment recommendations, drug stock management, drug distribution choices, and general acceptance of the need to prepare for the coming challenge of drug resistance [55–57]. These studies suggest that acceptability for changes in treatment policies is high and that novel drug management approaches will be feasible, however the final proof can only come from on-the-ground implementation occurring outside a research context. Adherence to ASAQ use will play an important factor in every strategy; reduced preference for ASAQ due to gastrointestinal intolerance or other symptoms will need to be monitored to determine if planned and realized ASAQ deployment coincide.

Third, in modeling analyses, ASAQ use will also have to be assessed alongside the use of amodiaquine in seasonal malaria chemoprophylaxis (SMC) programs which use a monthly combination of sulfadoxine-pyrimethamine and amodiaquine (SPAQ). Simultaneous use of ASAQ and SPAQ could increase the rate of AQ-resistance evolution. Although SMC has mostly been used in West Africa, it was recently initiated in northeastern Uganda [58] where malaria incidence is relatively seasonal. If SMC is expanded in Uganda, several million children may eventually be covered by the program, potentially receiving between 10 and 20 million courses of SPAQ, several million of which would be administered to parasite-positive children. This means that deployment of first-line ASAQ for symptomatic uncomplicated falciparum malaria in the public sector would entail using approximately 13 million ASAQ treatments in higher-parasitaemia symptomatic cases alongside several million SPAQ treatments for children with lower-parasitaemia asymptomatic infections. There is no simple way to determine the additional evolutionary pressure on amodiaquine resistance evolution generated by SPAQ use; thus, a separate modeling analysis with known SMC distribution data will need to be carried out. This separate analysis will also need to account for the importation of AQ-resistant genotypes from newly arriving populations from South Sudan where ASAQ is currently first-line therapy [59].

Fourth, an important unknown in Uganda is the 2025 allele frequency of circulating kelch variants whose district-level frequencies were last reported in 2022. As ACTs continue to be used in Uganda and across Africa, modeling analyses predict that genotype frequencies of kelch variants will increase in the near term. Our modeling analysis projects that the combined frequency of the *pfkelch13* 469Y and 675V alleles will be 0.91 (90% range: 0.88 – 0.94) in mid-2025, predicting that these slow-clearance phenotypes may already be near fixation in 14 or 15 districts in Uganda. However, we do not know whether these two genotypes account for all the phenotypes that cause long clearance times and potentially treatment failure; thus, it is impossible to know how accurate this projection will be on a per-allele or per-genotype basis. The strong selection pressure in our model is based on the high ACT coverage reported in Uganda (>80% in all MIS regions) and rapid movement and transportation of these parasites around the country. However, the most recent data from Uganda [60] indicate that at a regional level (looking at northern or eastern Uganda) each of these alleles was circulating between 0.05 and 0.30 allele frequency in 2024. The next iteration of country-level model calibration in Uganda will need to include validation metrics on the strength of selection pressure, the pace of evolution, the movement of parasites around the country, and an accounting (if possible) of all genotypes that are potentially being selected.

Finally, the procurement of all ACTs in Africa has been put in jeopardy in 2025 by the closing of the United States State Department’s President’s Malaria Initiative (PMI) which is estimated to leave a gap of approximately half of all ACTs procured in Africa [61]. The sudden loss of PMI programs and resources is projected to increase malaria deaths in Africa by approximately 100,000 in 2025 [61,62] and will undoubtedly have a large impact on country programs’ ability to prepare for the coming fight against spreading artemisinin resistance. The specific challenge of procuring more types of ACTs at a moment when financing mechanisms for these purchases have disappeared is unlikely to be easily met and was not modelled in the present analysis. The next iteration of treatment strategy evaluations should focus on cost, simplicity, and feasibility in case these funding cuts become permanent.

### Conclusions

As artemisinin-resistance response moves forward in Africa, continual monitoring and real-time data collection are essential. Therapeutic efficacy studies (TES) are a particularly important component of this effort, as this will be the only way to know when increasing *pfkelch13* allele frequencies and high-grade partner-drug resistance combine to drive treatment efficacies to low levels. Crucially, TES results must be accompanied by genotyping of polymorphisms known or strongly suspected to be associated with clinical drug-resistance phenotypes [45]. The population-level treatment strategies evaluated in this analysis act by favoring some genotypes over others, and the genotype information contained in TES studies is the most important way to know which genotypes are associated with failure and thus which genotypes should be targeted for reduction by a treatment strategy.

Adoption of novel treatment strategies this decade in the African countries where kelch variants are currently circulating will be critical for the rest of the continent as decisions are taken up country by country and as *pfkelch13* alleles arrive in new locations demanding new mitigation responses. In Uganda, the appropriate response – as projected from this modeling analysis – is to switch to ASAQ as a transition strategy and decide whether to proceed to a rotational approach, triple ACT adoption, or a variant of an MFT strategy. We recommend that this strategy change be instituted soon, as continued use of AL is projected to drive treatment failure counts higher every year. Once this challenge is met, the next step in the drug-resistance mitigation process – in Uganda and in all African countries that may be facing future treatment failure increases – will be to replace ACTs with non-artemisinin based combination therapies to begin the process, in the 2030s, of reversing the *pfkelch13* evolution that occurred in the late 2010s and 2020s.

## Data Availability

All simulation code and outputs are available at https://github.com/bonilab/Uganda-phase-1

## Supporting Information

See Supplementary Figures S1 to S6 and Supplementary Tables S1 to S3.

## Data and Code Availability

All relevant data and code can be found in the Github repository: https://github.com/bonilab/Uganda-phase-1

## Acknowledgements

TDN, RJZ, KTT, CCF, MFB, DMG were funded by National Institutes of Health grant NIAID R01AI153355 and Bill and Melinda Gates Foundation grant INV-005517. TDN, KTT, CCF, MFB were funded by National Institutes of Health grant NIAID R01AI153355 and Bill and Melinda Gates Foundation grant INV-056612. VA was funded by Bill and Melinda Gates Foundation grant INV-037316. GBR, BBA, MRK, JO are funded by US President’s Malaria Initiative prime contract 72061722C00003 and sub-contract 14070-IDRC-01. The study was additionally funded by the National Institutes of Health (R01AI075045, R01AI173557, U19AI089674, RO1AI117001, R01AI139179, and D43TW010526), the Medicines for Malaria Venture (RD/15/0001), and the Gates Foundation (INV-035751). Computations for this research were performed on the Pennsylvania State University’s Institute for Computational and Data Sciences’ Roar supercomputer and on Temple University’s College of Science and Technology Owl’s Nest Cluster.

## Declarations

PJR consulted for GSK in 2023 and Merck in 2024 on new antimalarial drug discovery.

## Author Contributions

**Conceptualization:** MFB, JO, BBA, MRK, GBR

**Formal Analysis:** TDN, RJZ

**Funding Acquisition:** MFB

**Investigation:** TDN, RJZ, MDC, CCF, VA, KTT, DMG

**Methodology:** TDN, RJZ, MDC, CCF, VA, KTT, DMG

**Supporting Data:** MDC, PJR, GBR, BBA, MRK, JO

**Software:** TDN, RJZ, KTT

**Supervision:** MFB, JO

**Visualization:** TDN, RJZ, CCF

**Writing – original draft**: MFB

**Writing – review & editing:** MFB, TDN, PJR, MDC.

## References

1. World Health Organization. World Malaria Report 2024.

2. World Health Organization. Artemisinin resistance and artemisinin-based combination therapy efficacy: status report. Geneva, Switzerland: World Health Organization; 2018. Report No.: WHO/CDS/GMP/2018.18. Available: https://iris.who.int/handle/10665/274362

3. Akala H, Opot B, Juma DW, Okoth RO, Mwalo M, Salim FA, et al. Detection of Twenty-Four Plasmodium Falciparum Kelch 13 Mutations Including C469Y, P553L, R561H, and A675V Across Kenya. Rochester, NY: Social Science Research Network; 2024. doi:10.2139/ssrn.5020665

4. van Wyk S, Seocharan I, Mlugu EM, Ayuen DS, Mategula D, Makhaza T, et al. The MARC SE-Africa Dashboard: Joining Forces to Counteract Emerging Antimalarial Resistance in South and East Africa. medRxiv; 2025. p. 2025.01.07.25320158. doi:10.1101/2025.01.07.25320158

5. Rosenthal PJ, Asua V, Conrad MD. Emergence, transmission dynamics and mechanisms of artemisinin partial resistance in malaria parasites in Africa. Nat Rev Microbiol. 2024;22: 373–384. doi:10.1038/s41579-024-01008-2

6. Dhorda M, Kaneko A, Komatsu R, KC A, Mshamu S, Gesase S, et al. Artemisinin-resistant malaria in Africa demands urgent action. Science. 2024;385: 252–254. doi:10.1126/science.adp5137

7. Rosenthal PJ, Asua V, Bailey JA, Conrad MD, Ishengoma DS, Kamya MR, et al. The emergence of artemisinin partial resistance in Africa: how do we respond? Lancet Infect Dis. 2024;24: e591–e600. doi:10.1016/S1473-3099(24)00141-5

8. Ishengoma DS, Gosling R, Martinez-Vega R, Beshir KB, Bailey JA, Chimumbwa J, et al. Urgent action is needed to confront artemisinin partial resistance in African malaria parasites. Nat Med. 2024;30: 1807–1808. doi:10.1038/d41591-024-00028-y

9. World Health Organization. Multiple first-line therapies as part of the response to antimalarial drug resistance: an implementation guide. Geneva, Switzerland; 2024 Nov. Available: https://who.int/publications/i/item/9789240103603

10. World Health Organization. Strategy to respond to antimalarial drug resistance in Africa. 2022.

11. Zupko RJ, Nguyen TD, Ngabonziza JCS, Kabera M, Li H, Tran TN-A, et al. Modeling policy interventions for slowing the spread of artemisinin-resistant pfkelch R561H mutations in Rwanda. Nat Med. 2023;10.1038/s41591-023-02551-w. Available: https://www.medrxiv.org/content/10.1101/2022.12.12.22283369v1

12. Boni MF, White NJ, Baird JK. The Community As the Patient in Malaria-Endemic Areas: Preempting Drug Resistance with Multiple First-Line Therapies. PLoS Med. 2016;13: e1001984. doi:10.1371/journal.pmed.1001984

13. Rueangweerayut R, Phyo AP, Uthaisin C, Poravuth Y, Binh TQ, Tinto H, et al. Pyronaridine–Artesunate versus Mefloquine plus Artesunate for Malaria. N Engl J Med. 2012;366: 1298–1309. doi:10.1056/NEJMoa1007125

14. Pryce J, Taylor M, Fox T, Hine P. Pyronaridine-artesunate for treating uncomplicated *Plasmodium falciparum* malaria. Cochrane Database Syst Rev. 2022 [cited 12 Jan 2025]. doi:10.1002/14651858.CD006404.pub4

15. Falade CO, Orimadegun AE, Olusola FI, Michael OS, Anjorin OE, Funwei RI, et al. Efficacy and safety of pyronaridine– artesunate versus artemether–lumefantrine in the treatment of acute uncomplicated malaria in children in South-West Nigeria: an open-labelled randomized controlled trial. Malar J. 2023;22: 154. doi:10.1186/s12936-023-04574-7

16. Alebachew M, Gelaye W, Abate MA, Sime H, Hailgiorgis H, Gidey B, et al. Therapeutic efficacy of pyronaridine-artesunate (Pyramax®) against uncomplicated Plasmodium falciparum infection at Hamusit Health Centre, Northwest Ethiopia. Malar J. 2023;22: 186. doi:10.1186/s12936-023-04618-y

17. Dimbu PR, Labuda S, Ferreira CM, Caquece F, André K, Pembele G, et al. Therapeutic response to four artemisinin-based combination therapies in Angola, 2021. Antimicrob Agents Chemother. 2024;68: e01525–23. doi:10.1128/aac.01525-23

18. Conrad MD, Asua V, Garg S, Giesbrecht D, Niaré K, Smith S, et al. Evolution of Partial Resistance to Artemisinins in Malaria Parasites in Uganda. N Engl J Med. 2023;389: 722–732. doi:10.1056/NEJMoa2211803

19. Zupko RJ, Nguyen TD, Somé AF, Tran TN-A, Gerardin J, Dudas P, et al. Long-term effects of increased adoption of artemisinin combination therapies in Burkina Faso. PLoS Glob Public Health. 2022;2: e0000111. doi:10.1371/journal.pgph.0000111

20. Humanitarian Data Exchange. Uganda - Subnational Administrative Boundaries - Humanitarian Data Exchange. [cited 5 Jan 2025]. Available: https://data.humdata.org/dataset/cod-ab-uga

21. USAID. DHS Program, Spatial Data Repository - Boundaries. Feb 2024. Available: https://spatialdata.dhsprogram.com/boundaries/#countryId=UG&view=table

22. WorldPop. Global 100m Population total adjusted to match the corresponding UNPD estimate. University of Southampton; 2020. doi:10.5258/SOTON/WP00660

23. Weiss DJ, Nelson A, Gibson HS, Temperley W, Peedell S, Lieber A, et al. A global map of travel time to cities to assess inequalities in accessibility in 2015. Nature. 2018;553: 333–336. doi:10.1038/nature25181

24. Wesolowski A, O’Meara WP, Eagle N, Tatem AJ, Buckee CO. Evaluating Spatial Interaction Models for Regional Mobility in Sub-Saharan Africa. PLOS Comput Biol. 2015;11: e1004267. doi:10.1371/journal.pcbi.1004267

25. Zupko RJ, Nguyen TD, Wesolowski A, Gerardin J, Boni MF. National-scale simulation of human movement in a spatially coupled individual-based model of malaria in Burkina Faso. Nat Sci Rep. 2023;13: 321. doi:10.1038/s41598-022-26878-5

26. ECMWF / Copernicus Climate Change Service. ERA5 Daily Aggregates - Latest Climate Reanalysis Produced by ECMWF / Copernicus Climate Change Service | Earth Engine Data Catalog. In: Google for Developers [Internet]. 2020 [cited 8 Jan 2025]. Available: https://developers.google.com/earth-engine/datasets/catalog/ECMWF_ERA5_DAILY

27. Malaria Atlas Project. Malaria Atlas Project | Data. [cited 8 Jan 2025]. Available: https://data.malariaatlas.org/trends?year=2020&metricGroup=Malaria&geographicLevel=admin0&metricSubcategory=Pf&metricType=rate&metricName=PR

28. Zupko RJ. Malaria Simulation (MaSim) Version: 4.1.7.1. In: ResearchGate [Internet]. 19 Dec 2024 [cited 8 Jan 2025]. doi:10.13140/RG.2.2.14091.20009

29. Uganda Ministry of Health National Malaria Control Division. Uganda Malaria Indicator Survey 2018-2019. 2020. Available: https://dhsprogram.com/pubs/pdf/MIS34/MIS34.pdf

30. World Health Organization. World Malaria Report 2024. [cited 21 Mar 2025]. Available: https://www.who.int/teams/global-malaria-programme/reports/world-malaria-report-2024

31. Balikagala B, Fukuda N, Ikeda M, Katuro OT, Tachibana S-I, Yamauchi M, et al. Evidence of Artemisinin-Resistant Malaria in Africa. N Engl J Med. 2021;385: 1163–1171. doi:10.1056/NEJMoa2101746

32. Ikeda M, Kaneko M, Tachibana S-I, Balikagala B, Sakurai-Yatsushiro M, Yatsushiro S, et al. Artemisinin-Resistant Plasmodium falciparum with High Survival Rates, Uganda, 2014–2016 - Volume 24, Number 4—April 2018 - Emerging Infectious Diseases journal - CDC. [cited 10 Jan 2025]. doi:10.3201/eid2404.170141

33. Asua V, Vinden J, Conrad MD, Legac J, Kigozi SP, Kamya MR, et al. Changing Molecular Markers of Antimalarial Drug Sensitivity across Uganda. Antimicrob Agents Chemother. 2019;63: e01818–18. doi:10.1128/AAC.01818-18

34. Tumwebaze PK, Conrad MD, Okitwi M, Orena S, Byaruhanga O, Katairo T, et al. Decreased susceptibility of Plasmodium falciparum to both dihydroartemisinin and lumefantrine in northern Uganda. Nat Commun. 2022;13: 6353. doi:10.1038/s41467-022-33873-x

35. Ansel J, Yang E, He H, Gimelshein N, Jain A, Voznesensky M, et al. PyTorch 2: Faster Machine Learning Through Dynamic Python Bytecode Transformation and Graph Compilation. Proceedings of the 29th ACM International Conference on Architectural Support for Programming Languages and Operating Systems, Volume 2. New York, NY, USA: Association for Computing Machinery; 2024. pp. 929–947. doi:10.1145/3620665.3640366

36. Reiker T, Golumbeanu M, Shattock A, Burgert L, Smith TA, Filippi S, et al. Emulator-based Bayesian optimization for efficient multi-objective calibration of an individual-based model of malaria. Nat Commun. 2021;12: 7212. doi:10.1038/s41467-021-27486-z

37. Akiba T, Sano S, Yanase T, Ohta T, Koyama M. Optuna: A Next-generation Hyperparameter Optimization Framework. Proceedings of the 25th ACM SIGKDD International Conference on Knowledge Discovery & Data Mining. New York, NY, USA: Association for Computing Machinery; 2019. pp. 2623–2631. doi:10.1145/3292500.3330701

38. Watson OJ, Gao B, Nguyen TD, Tran TN-A, Penny MA, Smith DL, et al. Pre-existing partner-drug resistance facilitates the emergence and spread of artemisinin resistance: a consensus modelling study. Lancet Microbe. 2022. pp. 701–710. doi:10.1101/2021.04.08.437876

39. Pongtavornpinyo W, Hastings IM, Dondorp A, White LJ, Maude RJ, Saralamba S, et al. Probability of emergence of antimalarial resistance in different stages of the parasite life cycle. Evol Appl. 2009;2: 52–61. doi:10.1111/j.1752-4571.2008.00067.x

40. Asua V, Conrad MD, Aydemir O, Duvalsaint M, Legac J, Duarte E, et al. Changing Prevalence of Potential Mediators of Aminoquinoline, Antifolate, and Artemisinin Resistance Across Uganda. J Infect Dis. 2021;223: 985–994. doi:10.1093/infdis/jiaa687

41. Witkowski B, Duru V, Khim N, Ross LS, Saintpierre B, Beghain J, et al. A surrogate marker of piperaquine-resistant Plasmodium falciparum malaria: a phenotype–genotype association study. Lancet Infect Dis. 2017;17: 174–183. doi:10.1016/S1473-3099(16)30415-7

42. Uganda Ministry of Health. Strategy to Respond to Antimalarial Drug Resistance in Uganda. Kampala, Uganda: [private]; 2025.

43. Boni MF, Smith DL, Laxminarayan R. Benefits of using multiple first-line therapies against malaria. Proc Natl Acad Sci USA. 2008;105: 14216–14221. doi:10.1073/pnas.0804628105

44. Nguyen TD, Gao B, Amaratunga C, Dhorda M, Tran TN-A, White NJ, et al. Preventing antimalarial drug resistance with triple artemisinin-based combination therapies. Nat Commun. 2023;14: 4568. doi:10.1038/s41467-023-39914-3

45. Tran KT, Nguyen TD, Weissman DB, Li EZ, Mok S, Small-Saunders JL, et al. Effects of recombination on multi-drug resistance evolution in Plasmodium falciparum malaria. bioRxiv; 2024. p. 2024.12.18.628924. doi:10.1101/2024.12.18.628924

46. Mavoko HM, Nabasumba C, da Luz RI, Tinto H, D’Alessandro U, Kambugu A, et al. Efficacy and safety of re-treatment with the same artemisinin-based combination treatment (ACT) compared with an alternative ACT and quinine plus clindamycin after failure of first-line recommended ACT (QUINACT): a bicentre, open-label, phase 3, randomised controlled trial. Lancet Glob Health. 2017;5: e60–e68. doi:10.1016/S2214-109X(16)30236-4

47. Ebong C, Sserwanga A, Namuganga JF, Kapisi J, Mpimbaza A, Gonahasa S, et al. Efficacy and safety of artemether-lumefantrine and dihydroartemisinin-piperaquine for the treatment of uncomplicated Plasmodium falciparum malaria and prevalence of molecular markers associated with artemisinin and partner drug resistance in Uganda. Malar J. 2021;20: 484. doi:10.1186/s12936-021-04021-5

48. Agaba BB, Travis J, Smith D, Rugera SP, Zalwango MG, Opigo J, et al. Emerging threat of artemisinin partial resistance markers (pfk13 mutations) in Plasmodium falciparum parasite populations in multiple geographical locations in high transmission regions of Uganda. Malar J. 2024;23: 330. doi:10.1186/s12936-024-05158-9

49. Pryce J, Taylor M, Fox T, Hine P. Pyronaridine-artesunate for treating uncomplicated *Plasmodium falciparum* malaria. Cochrane Database Syst Rev. 2022 [cited 25 Sep 2024]. doi:10.1002/14651858.CD006404.pub4

50. van der Pluijm RW, Imwong M, Chau NH, Hoa NT, Thuy-Nhien NT, Thanh NV, et al. Determinants of dihydroartemisinin-piperaquine treatment failure in Plasmodium falciparum malaria in Cambodia, Thailand, and Vietnam: a prospective clinical, pharmacological, and genetic study. Lancet Infect Dis. 2019;19: 952–961. doi:10.1016/S1473-3099(19)30391-3

51. Mok S, Yeo T, Hong D, Shears MJ, Ross LS, Ward KE, et al. Mapping the genomic landscape of multidrug resistance in Plasmodium falciparum and its impact on parasite fitness. Sci Adv. 2023;9: eadi2364. doi:10.1126/sciadv.adi2364

52. Small-Saunders JL, Hagenah LM, Wicht KJ, Dhingra SK, Deni I, Kim J, et al. Evidence for the early emergence of piperaquine-resistant Plasmodium falciparum malaria and modeling strategies to mitigate resistance. PLOS Pathog. 2022;18: e1010278. doi:10.1371/journal.ppat.1010278

53. Ross LS, Dhingra SK, Mok S, Yeo T, Wicht KJ, Kümpornsin K, et al. Emerging Southeast Asian PfCRT mutations confer Plasmodium falciparum resistance to the first-line antimalarial piperaquine. Nat Commun. 2018;9: 3314. doi:10.1038/s41467-018-05652-0

54. Dhingra SK, Small-Saunders JL, Ménard D, Fidock DA. Plasmodium falciparum resistance to piperaquine driven by PfCRT. Lancet Infect Dis. 2019;19: 1168–1169. doi:10.1016/S1473-3099(19)30543-2

55. Kaboré JMT, Siribié M, Hien D, Soulama I, Barry N, Baguiya A, et al. Feasibility and Acceptability of a Strategy Deploying Multiple First-Line Artemisinin-Based Combination Therapies for Uncomplicated Malaria in the Health District of Kaya, Burkina Faso. Trop Med Infect Dis. 2023;8: 195. doi:10.3390/tropicalmed8040195

56. Hien D, Kaboré JMT, Siribié M, Soulama I, Barry N, Baguiya A, et al. Stakeholder perceptions on the deployment of multiple first-line therapies for uncomplicated malaria: a qualitative study in the health district of Kaya, Burkina Faso. Malar J. 2022;21: 202. doi:10.1186/s12936-022-04225-3

57. Chege T, Cole A, Aman R, Githuka G, Muga R, Kokwaro G. Health System Challenges Associated with Deployment of Multiple First Line Treatment for Uncomplicated Malaria: A Pilot Study in a Malaria-Endemic Region of Kenya. SSRN; 2024. doi:10.2139/ssrn.4756459

58. Nuwa A, Baker K, Kajubi R, Nnaji CA, Theiss-Nyland K, Odongo M, et al. Effectiveness of sulfadoxine–pyrimethamine plus amodiaquine and dihydroartemisinin–piperaquine for seasonal malaria chemoprevention in Uganda: a three-arm, open-label, non-inferiority and superiority, cluster-randomised, controlled trial. Lancet Infect Dis. 2025;0. doi:10.1016/S1473-3099(24)00746-1

59. Tukwasibwe S, Garg S, Katairo T, Asua V, Kagurusi BA, Mboowa G, et al. Varied Prevalence of Antimalarial Drug Resistance Markers in Different Populations of Newly Arrived Refugees in Uganda. J Infect Dis. 2024;230: 497–504. doi:10.1093/infdis/jiae288

60. Okitwi M, Orena S, Tumwebaze PK, Katairo T, Taremwa Y, Byaruhanga O, et al. Changes in susceptibility of Plasmodium falciparum to antimalarial drugs in Uganda over time: 2019-2024. medRxiv; 2025. p. 2024.12.31.24319821. doi:10.1101/2024.12.31.24319821

61. Symons TL, Lubinda J, McPhail M, Saddler A, Berg M van den, Baggen H, et al. Estimating the potential malaria morbidity and mortality avertable by the President’s Malaria Initiative in 2025: a geospatial modelling analysis. medRxiv; 2025. p. 2025.02.28.25323072. doi:10.1101/2025.02.28.25323072

62. Winskill P, Haile L, Rybal-Pesantez S, Simmons O, Topazian H, Okell, Lucy. Rapid response modelled estimates of the effect of the US global aid freeze on President’s Malaria Initiative impact in sub-Saharan Africa. Imperial College London; 2025. 10.25561/118190.

