## Supplementary Figs S1-S6 and Table S1 for "Slowing the spread of treatment failure to artemisinin-based combination therapies in Uganda"

### Supplementary Materials Table of Contents

#### S1. Supplementary Figures

- **Figure S1.** Construction of the rainfall-derived seasonal transmission function used in the Uganda malaria simulation model.
- **Figure S2.** Monthly treatment failures under 1-year cycling strategies (2025–2031)
- **Figure S3.** Monthly treatment failures under 3-year cycling strategies (2025–2031)
- **Figure S4.** Travel time to nearest city (gravity model input)
- **Figure S5.** Overlay of *pfkelch13* allele frequencies and modelled travel density
- **Figure S6.** Simulated travel density surface used for validation

#### S2. Supplementary Tables

- **Table S1.** Summary of treatment failures, failure rates, and pfkelch13 allele frequencies across all evaluated strategies with percent change from status quo
- **Table S2.** *PfPR* calibration inputs by district, showing Plasmodium falciparum prevalence estimates used for model fitting. (Supp Tables – S2 and S3.xlsx)
- **Table S3.** Compilation of *pfkelch13* C469Y and A675V allele frequencies by district and year, used for model calibration of resistance allele spread in Uganda. (Supp Tables – S2 and S3.xlsx)

### S1. Supplementary Figures

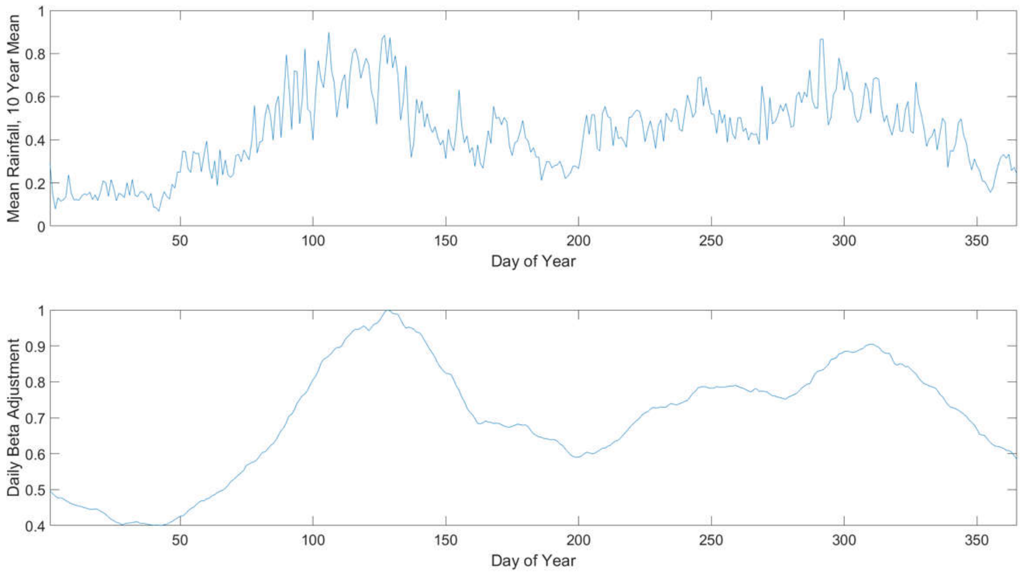


**Supplementary Figure S1.** Construction of the rainfall-derived seasonal transmission function used in the Uganda malaria simulation model. Top panel shows mean daily rainfall across Uganda, averaged over the period 2009–2019 from ERA5. Bottom panel shows daily scaling factor applied to the transmission rate parameter $\beta$ (t) representing seasonal variation in the Anopheles biting rate. This adjustment curve was smoothed and shifted forward by 11 days to reflect the delay between rainfall and peak mosquito abundance. The same within-year $\beta$ (t) profile was applied uniformly across all spatial locations in the model.

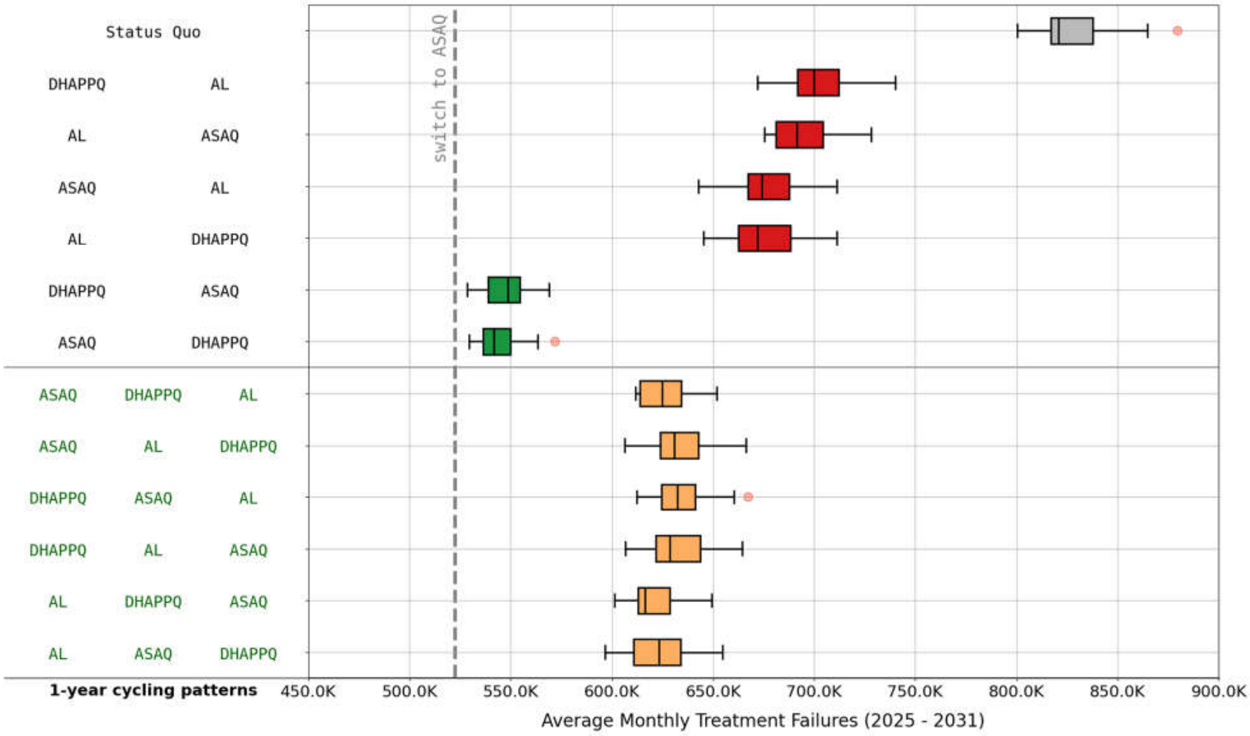


**Figure S2.** Average Monthly Treatment Failures (2025 - 2031) under Different 1-Year Cycling Strategies. This figure replicates Figure 5 from the main text with 1-year cycling policies. The labels on the left show the order (left-to-right) of therapies used in each cycling regimen.

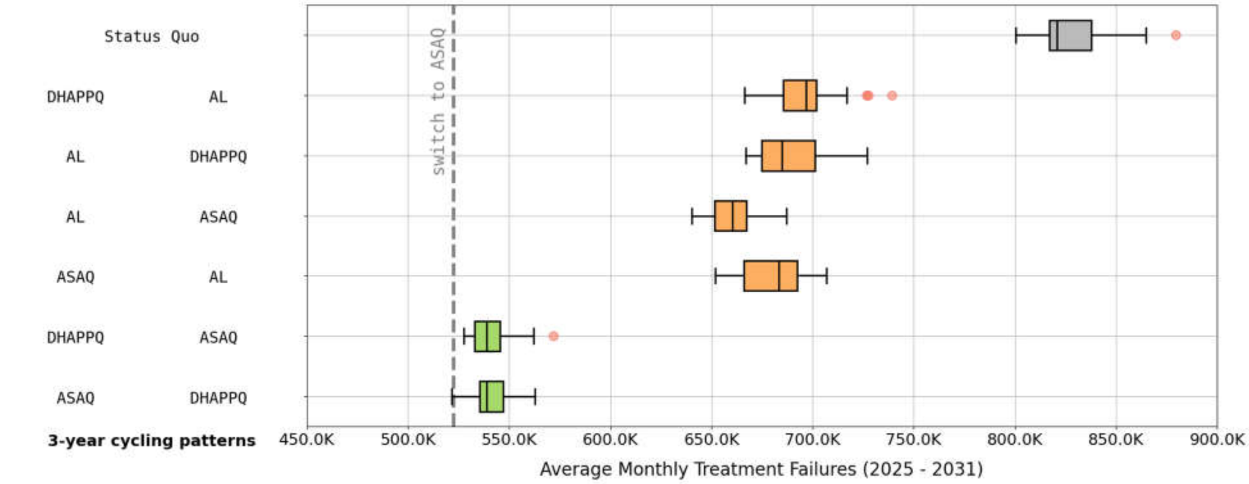


**Figure S3.** Average Monthly Treatment Failures (2025 - 2031) under Different 3-Year Cycling Strategies. This figure replicates Figure 5 from the main text with 3-year cycling policies. The labels on the left show the order (left-to-right) of therapies used in each cycling regimen.

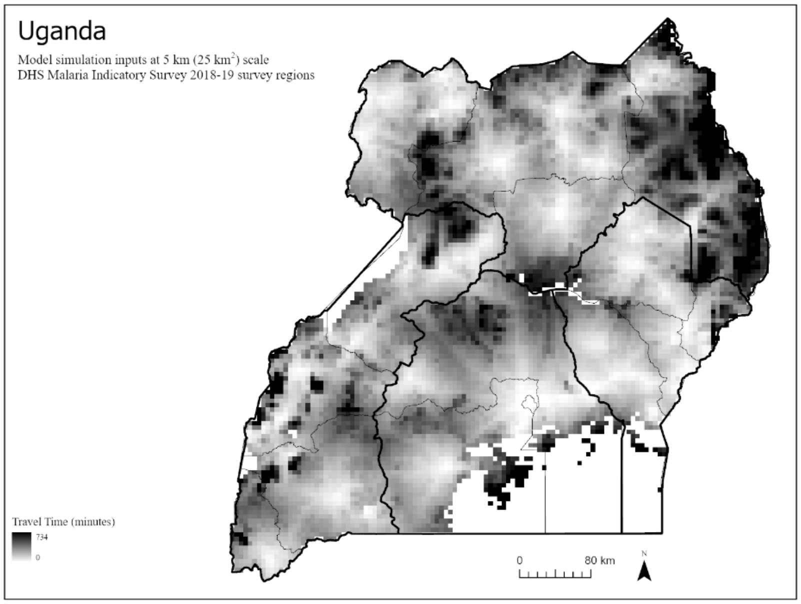


**Figure S4.** Estimated travel time to the nearest city across Uganda, aggregated to 25 $km^2$ resolution. These values represent the travel friction surface used as input to a gravity-based human movement model. Higher travel time values (darker shades) indicate more isolated regions with lower accessibility to urban centers.

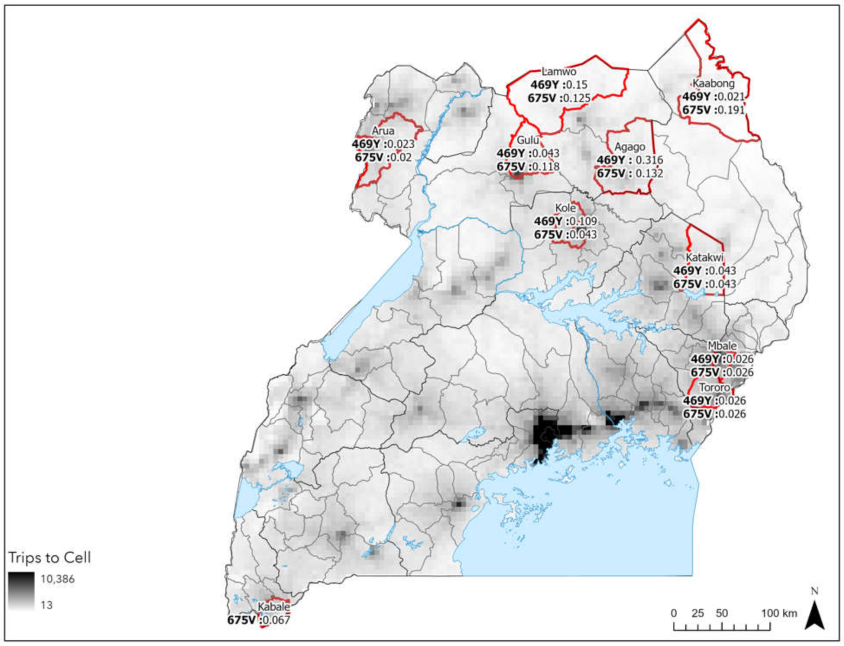


**Figure S5.** Spatial overlay of observed pfkelch13 allele frequencies (469Y and 675V) and simulated travel density from the gravity model. Red boundaries indicate districts with field-verified presence of kelch variants. Background shading represents the number of simulated trips per pixel, highlighting connectivity to and from regions with known resistance.

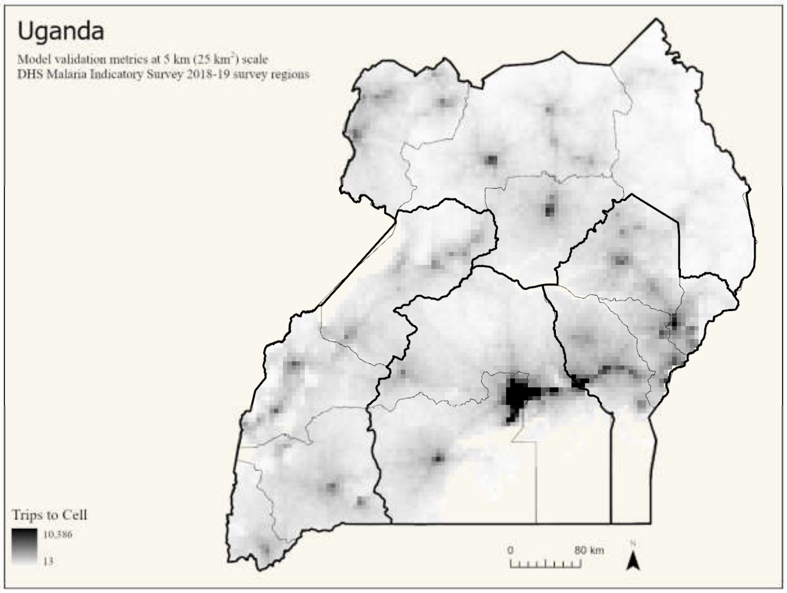


**Figure S6.** Simulated travel density surface at 25 km$^2$ resolution, representing the cumulative number of trips arriving at each pixel across all model-runs. This surface was used for validation of the human movement model and to assess spatial connectivity across epidemiologically relevant districts.

#### S2. Supplementary Tables

| Strategy | Annual TF (median) | Monthly TF (median) | %Δ vs Status Quo | TF Rate 2025-26 (median) | TF Rate 2030-31 (median) |
|---|---|---|---|---|---|
| *Continued AL use (status quo)* | 9.9M | 820,898 | 0.00% | 26.20% | 33.10% |
| *Switch to ASAQ* | 6.3M | 522,456 | -36.40% | 19.20% | 19.70% |
| *Switch to ASAQ* | 8.7M | 728,884 | -11.20% | 19.30% | 37.60% |
| *2-year cycling: AL → ASAQ* | 8.7M | 722,369 | -12.00% | 26.00% | 32.00% |
| *2-year cycling: AL → DHA-PPQ* | 8.8M | 732,470 | -10.80% | 26.30% | 32.60% |
| *2-year cycling: ASAQ → AL* | 7.4M | 619,887 | -24.50% | 19.20% | 20.10% |
| *2-year cycling: ASAQ → DHA-PPQ* | 6.2M | 520,568 | -36.60% | 19.10% | 19.70% |
| *2-year cycling: DHA-PPQ → AL* | 8.3M | 691,259 | -15.80% | 19.20% | 30.70% |
| *2-year cycling: DHA-PPQ → ASAQ* | 7.0M | 587,119 | -28.50% | 19.10% | 30.20% |
| *2-year cycling: AL → ASAQ → DHA-PPQ* | 7.3M | 607,906 | -25.90% | 26.40% | 21.60% |
| *2-year cycling: AL → DHA-PPQ → ASAQ* | 7.4M | 618,165 | -24.70% | 26.70% | 20.40% |
| *2-year cycling: ASAQ → AL → DHA-PPQ* | 7.5M | 626,150 | -23.70% | 19.40% | 21.70% |
| *2-year cycling: ASAQ → DHA-PPQ → AL* | 7.5M | 626,222 | -23.70% | 19.00% | 30.90% |
| *2-year cycling: DHA-PPQ → AL → ASAQ* | 7.5M | 620,887 | -24.40% | 19.10% | 20.00% |
| *2-year cycling: DHA-PPQ → ASAQ → AL* | 7.5M | 627,094 | -23.60% | 19.10% | 30.90% |
| *MFT: 25% AL, 75% ASAQ* | 7.1M | 595,284 | -27.50% | 21.00% | 22.80% |
| *MFT: 25% AL, 75% DHA-PPQ* | 8.4M | 696,822 | -15.10% | 20.90% | 33.50% |
| *MFT: 50% AL, 50% ASAQ* | 8.0M | 667,406 | -18.70% | 22.60% | 26.00% |
| *MFT: 50% AL, 50% DHA-PPQ* | 8.4M | 698,098 | -15.00% | 22.90% | 30.10% |
| *MFT: 75% AL, 25% ASAQ* | 9.0M | 750,430 | -8.60% | 24.70% | 29.70% |
| *MFT: 75% AL, 25% DHA-PPQ* | 9.0M | 748,395 | -8.80% | 24.70% | 30.30% |
| *MFT: 25% ASAQ, 75% DHA-PPQ* | 7.6M | 630,079 | -23.20% | 19.30% | 30.70% |
| *MFT: 50% ASAQ, 50% DHA-PPQ* | 6.6M | 550,783 | -32.90% | 19.00% | 23.70% |
| *MFT: 75% ASAQ, 25% DHA-PPQ* | 6.3M | 521,649 | -36.50% | 19.20% | 20.20% |
| *MFT: 33% AL, 33% ASAQ, 33% DHA-PPQ* | 7.5M | 624,462 | -23.90% | 21.50% | 25.10% |
| *MFT: 5% AL, 47.5% ASAQ, 47.5% DHA-PPQ* | 7.1M | 592,199 | -27.90% | 20.70% | 23.80% |
| *DHA-PPQ for 4 years, then ALAQ* | 6.8M | 568,566 | -30.70% | 19.20% | 17.70% |
| *ASAQ for 4 years, then ALAQ* | 6.0M | 503,067 | -38.70% | 19.00% | 17.80% |
| *AL for 4 years, then ALAQ* | 8.5M | 706,494 | -13.90% | 26.90% | 19.20% |
| *2 years AL → 2 years ASAQ, then ALAQ* | 7.2M | 597,294 | -27.20% | 26.50% | 18.70% |
| *2 years AL → 2 years DHA-PPQ, then ALAQ* | 7.1M | 595,683 | -27.40% | 26.40% | 18.70% |
| *2 years ASAQ → 2 years AL, then ALAQ* | 7.3M | 609,280 | -25.80% | 19.20% | 18.70% |
| *2 years ASAQ → 2 years DHA-PPQ, then ALAQ* | 6.1M | 508,574 | -38.00% | 19.30% | 18.20% |
| *2 years DHA-PPQ → 2 years AL, then ALAQ* | 7.3M | 611,490 | -25.50% | 19.30% | 18.60% |
| *2 years DHA-PPQ → 2 years ASAQ, then ALAQ* | 6.1M | 506,830 | -38.30% | 19.20% | 18.00% |

| | | | | | |
|---|---|---|---|---|---|
| *Immediate ALAQ deployment* | 5.7M | 477,929 | -41.80% | 17.60% | 18.20% |
| *3-year cycling: AL → ASAQ* | 7.9M | 660,325 | -19.60% | 26.40% | 20.40% |
| *3-year cycling: AL → DHA-PPQ* | 8.2M | 684,893 | -16.60% | 26.70% | 25.60% |
| *3-year cycling: ASAQ → AL* | 8.2M | 683,255 | -16.80% | 19.30% | 31.70% |
| *3-year cycling: ASAQ → DHA-PPQ* | 6.5M | 538,964 | -34.30% | 19.20% | 24.70% |
| *3-year cycling: DHA-PPQ → AL* | 8.4M | 696,892 | -15.10% | 19.10% | 31.50% |
| *3-year cycling: DHA-PPQ → ASAQ* | 6.5M | 538,910 | -34.40% | 19.10% | 19.50% |
| *1-year cycling: AL → DHA-PPQ* | 8.1M | 671,989 | -18.10% | 26.20% | 22.60% |
| *1-year cycling: AL → ASAQ* | 8.3M | 691,637 | -15.70% | 26.20% | 27.10% |
| *1-year cycling: ASAQ → AL* | 8.1M | 674,166 | -17.90% | 19.10% | 29.60% |
| *1-year cycling: ASAQ → DHA-PPQ* | 6.5M | 541,889 | -34.00% | 19.20% | 24.30% |
| *1-year cycling: DHA-PPQ → AL* | 8.4M | 699,887 | -14.70% | 19.10% | 31.00% |
| *1-year cycling: DHA-PPQ → ASAQ* | 6.6M | 548,783 | -33.10% | 19.40% | 21.10% |
| *1-year cycling: AL → ASAQ → DHA-PPQ* | 7.5M | 623,339 | -24.10% | 26.90% | 21.60% |
| *1-year cycling: AL → DHA-PPQ → ASAQ* | 7.4M | 616,581 | -24.90% | 26.30% | 20.30% |
| *1-year cycling: ASAQ → AL → DHA-PPQ* | 7.6M | 630,946 | -23.10% | 19.20% | 24.00% |
| *1-year cycling: ASAQ → DHA-PPQ → AL* | 7.5M | 625,081 | -23.90% | 18.80% | 29.60% |
| *1-year cycling: DHA-PPQ → AL → ASAQ* | 7.5M | 628,812 | -23.40% | 19.20% | 22.40% |
| *1-year cycling: DHA-PPQ → ASAQ → AL* | 7.6M | 632,607 | -22.90% | 19.30% | 29.50% |

**Table S1.** Summary of treatment outcomes and resistance indicators across evaluated antimalarial deployment strategies. This table reports key simulation outcomes for each treatment strategy, including annual and monthly treatment failures and treatment failure rates in early and late periods (2025–2026 and 2030–2031). Percentage changes are shown relative to the status quo (continued AL use) as a baseline comparator.
